# Multicountry validation of a proteomic host-response signature associated with tuberculosis disease severity

**DOI:** 10.1101/2025.04.02.25324687

**Authors:** Zaynab Mousavian, Elin Folkesson, Xubin Zheng, Linn Kleberg, Maia Serene Gower, Laura Böttcher, Gina Gualano, Fabrizio Palmieri, Valentina Vanini, Gilda Cuzzi, Carolina Sousa Silva, Margarida Correia-Neves, Delia Goletti, Judith Bruchfeld, Gunilla Källenius, Christopher Sundling

## Abstract

**Introduction:** Non-sputum-based host-response biomarkers are needed to improve detection of tuberculosis (TB) across heterogeneous clinical presentations, particularly in individuals with negative sputum results. We previously identified a 12-marker plasma protein signature associated with TB disease severity. Here, we evaluated the reproducibility and generalizability of this signature across independent cohorts from Sweden and Italy, and assessed whether the panel could be reduced while maintaining diagnostic performance.

**Methods:** We analyzed 387 plasma samples from 314 participants, including individuals with TB disease, TB infection, and other non-TB diseases (non-TBD). Plasma proteomic profiling was used to validate enrichment of the original signature across cohorts with differing clinical spectra. The 12-marker signature was reduced to 6-protein (CDCP1, VEGFA, IFN-γ, CXCL9, IL-6, MCP-3) and 4-protein (CDCP1, IL-6, IFN-γ, CXCL9) signatures, and performance was compared with that of ten previously published protein signatures for TB disease.

**Results:** Both 6-marker and 4-marker signatures demonstrated enrichment in TB disease across cohorts and showed comparable performance to the original 12-marker panel and ten published protein signatures. At a fixed specificity of 70%, sensitivity for distinguishing TB disease from TB infection was 84% in the combined cohort and 94% in the Italian cohort. Sensitivity for distinguishing TB disease from non-TBD was 81% at the same specificity. Signature enrichment was consistent across heparin and EDTA plasma samples, and elevated levels of key cytokines were confirmed using independent bead-based platforms.

**Conclusions:** Together, these findings demonstrate robust external validation of a condensed host-response protein signature capturing TB-associated inflammatory states across populations and sample collection methods. The 6- and 4-protein signatures warrant further evaluation in translational studies of non–sputum-based diagnostics for pulmonary and extrapulmonary TB.

## Introduction

Tuberculosis (TB) disease, caused by Mycobacterium tuberculosis (*Mtb*) is the leading cause of mortality from an infectious disease, responsible for approximately 1.25 million deaths annually (1). It is also the leading cause of death among people living with HIV and a major contributor to antimicrobial resistance-related fatalities. It is estimated that each person with untreated pulmonary TB disease can infect 10 to 15 individuals per year (2). To reduce disease transmission and given the difficulty of diagnosing individuals with extrapulmonary (3, 4) and pediatric TB (2), there is an urgent need to develop better screening and diagnostic tests for TB disease (5), with particular emphasis on non-sputum-based biomarkers, as described by the World Health Organization (WHO) (6).

With advancements in omics technologies, new candidate biomarkers for diagnosis of TB disease are being evaluated (7, 8). In a recent review of 92 studies evaluating protein biomarkers for TB disease (9), we highlighted how methodology and protein abundance strongly impacted which proteins, individual or combined into signatures, were identified. However, most of the proteomic signatures have not been further validated in independent TB disease cohorts and, more importantly, in the context of other relevant non-TB diseases.

The primary aim of this study was to validate the accuracy and generalizability of a previously identified 12-marker plasma protein signature associated with TB disease severity (10) in independent cohorts from Sweden and Italy. We assessed signature accuracy across individuals with TB disease, TB infection, and clinically relevant non_-_TB diseases, and evaluated robustness across different plasma anticoagulants. As a secondary aim, we investigated whether the original signature could be condensed into smaller protein panels while maintaining performance. We then compared these signatures with other previously published proteomic signatures that overlapped with the current analytical platform.

## Results

### Clinical characteristics of the study participants

A total of 387 plasma samples from 314 individuals were included, divided into two subgroups: discovery or training (n=45) and validation or testing (n=342). The Swedish training cohort, as previously described in our biomarker discovery study (10) comprised 45 individuals: TB disease (n=21), TB infection (n=14), and IGRA-negative controls (n=10). In the current study, we used the discovery cohort for model training and signature refinement, while the validation cohort was employed to assess the accuracy of the identified markers in a new, independent cohort. Samples for the validation cohort were collected at two sites: Sweden and Italy. The Swedish cohort (n=136) consisted of individuals with pulmonary and extrapulmonary TB disease (n=30), TB infection (n=63), other non-TB diseases initially investigated for TB disease at the Karolinska University Hospital (n=26), and IGRA-negative controls (n=17). Heparin plasma was collected for all individuals, while EDTA plasma was collected from a subset of donors with TB disease (n=25), TB infection (n=23), and other non-TB diseases (n=25). The Italian cohort consisted of four groups: individuals with pulmonary TB disease (n=31), TB infection (n=56), other relevant non-TB diseases (n=26) and IGRA-negative controls (n=20). A schematic representation of the study plan and general cohort information is included in **Figure 1** with demographic and clinical information about the individual cohorts in **Supplementary Tables 1-3.**

**Figure 1.**
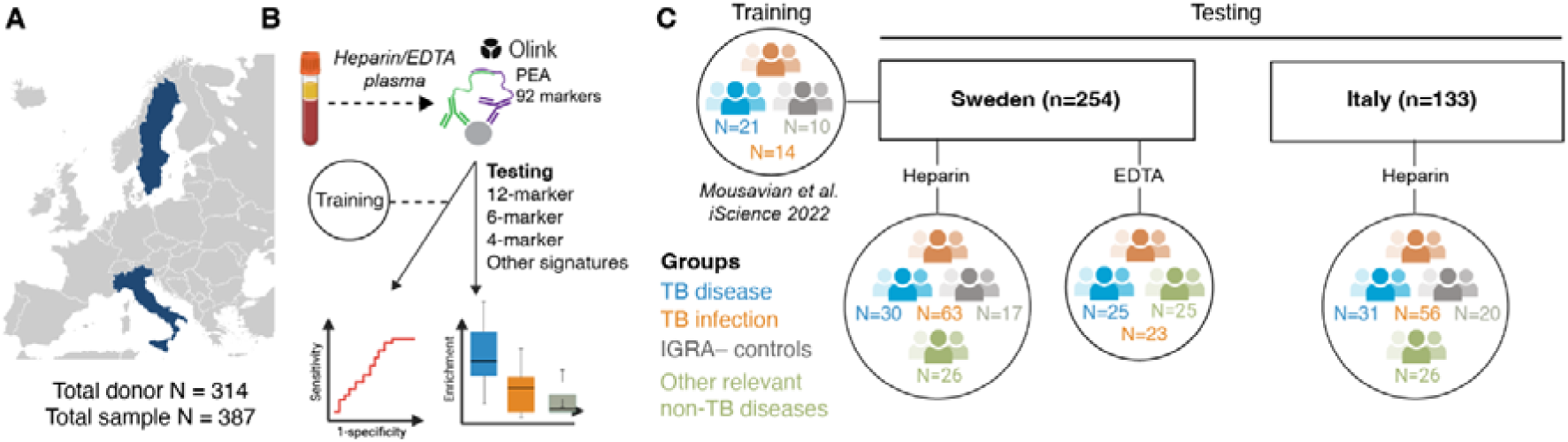
A schematic representation of the study plan and cohorts. (**A**) Study participants (n=314) were recruited from Sweden and Italy. (**B**) Blood samples were collected in heparin (n=314) and for a subset of donors in EDTA plasma (n=73) tubes and the Olink Target 96 Inflammation panel was used for measuring 92 proteins. 12-, 6-, and 4-marker or/and other published signatures for TB disease were evaluated for enrichment in the samples. The accuracy of each signature in identification of TB disease was assessed using receiver operating characteristics (ROC) analysis. (**C**) Each signature was first used to train a model on a repurposed dataset and then each model was tested in combined and separate Swedish and Italian validation cohorts using heparin plasma. When assessing accuracy in detecting TB disease versus other relevant non-TB diseases, both Swedish and Italian datasets were combined. Half of the dataset was used for training and the other half for testing. Additionally, signature enrichment and correlations were assessed between heparin and EDTA plasma in selected Swedish samples. **Abbreviations:** TB: tuberculosis; IGRA: Interferon-γ release assay; PEA: Proximity Extension Assay.

### Accuracy of the 12-marker signature in validation cohorts

We identified a 12-marker signature associated with TB disease, consisting of the proteins IFN-γ, IL-6, CDCP1, CXCL9, MMP-1, MCP-3, CCL19, CD40, VEGFA, IL-7, IL-12B (subunit p40), and PD-L1. To evaluate the accuracy of this signature in the new independent cohorts, we first used single-sample gene set enrichment analysis (ssGSEA) to determine whether the signature was enriched in individuals with TB disease compared to TB infection and IGRA-negative controls. The 12-marker signature was significantly enriched in the TB disease group compared to the two other groups (**Figure 2**). Sub-analysis of the Swedish and Italian validation cohorts separately showed that the enrichment score was lower for the Swedish samples. There was no significant difference between pulmonary TB and extra pulmonary TB disease that could explain this difference (**Supplementary Figure 1**). However, the enrichment score in individuals with TB disease, found early in disease progression through contact tracing, was very low (**Supplementary** Figure 1). Additionally, 40% of the Swedish donors with TB disease had no general TB symptoms. (fever, night sweats, loss of appetite, and weight loss), and 60% of the individuals with pulmonary TB were smear negative (compared with 16% in the Italian cohort), indicating more donors with mild disease (or asymptomatic TB) in the Swedish cohort (**Supplementary Table 1-3**).

**Figure 2.**
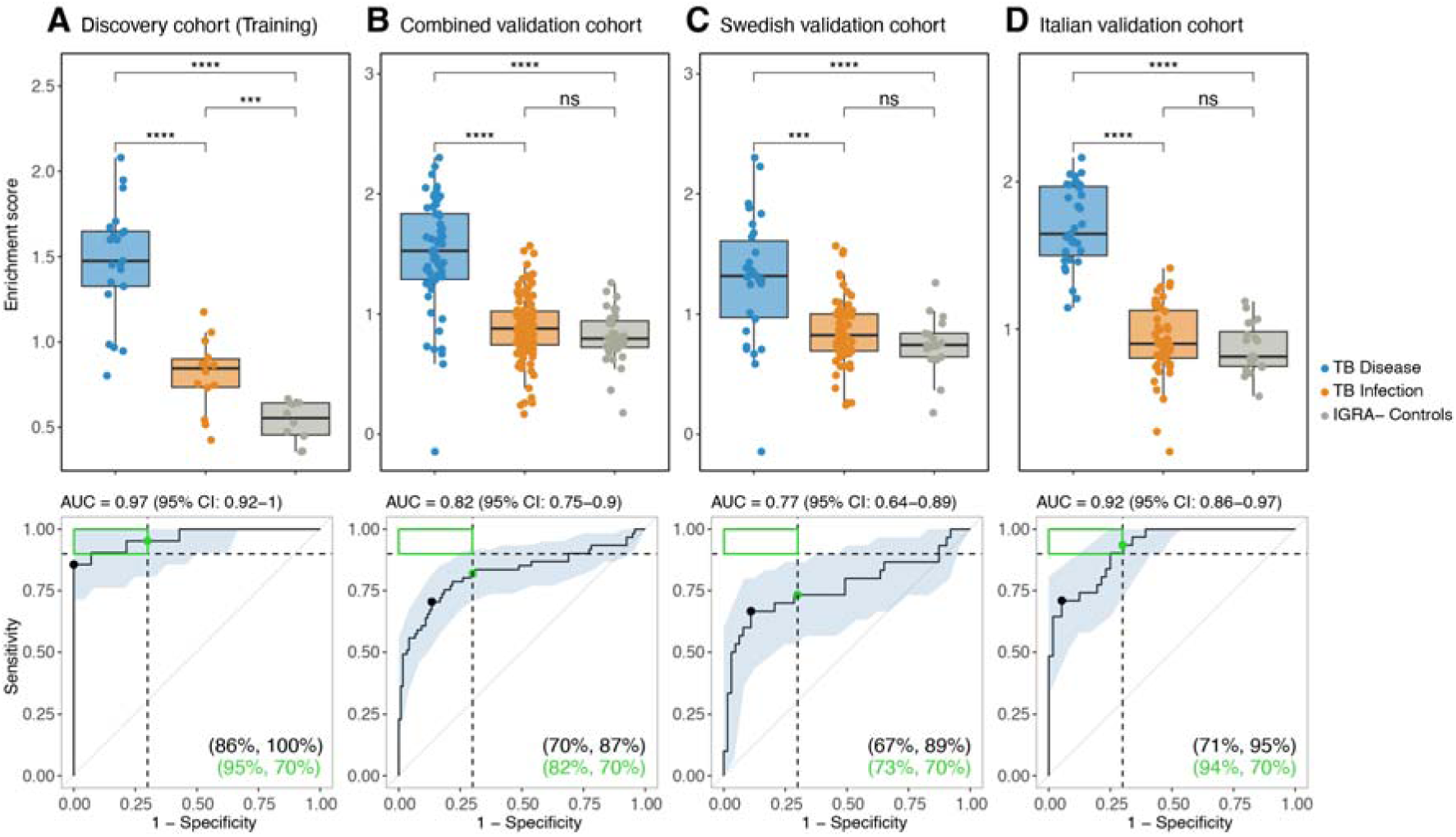
Accuracy of the 12-marker signature for TB disease compared to TB infection. The top row indicates a single-sample gene set enrichment analysis while the bottom row indicates ROC analysis, which was performed to distinguish TB disease from TB infection. The shaded area indicates a 95% confidence interval of the ROC curve, while the green box in the upper left corner indicates the minimum requirement for a triage test (sensitivity 90% and specificity 70%). The sensitivity at a fixed specificity of 70% for TB disease detection is indicated by a green point and the Youden index with a black point, percentage in the bottom-right corner of the graph. The area under the ROC curve (AUC) with 95% confidence interval is indicated above each graph. (**A**) Training cohort, (**B**) combined validation cohort, (**C**) Swedish validation cohort, and (**D**) Italian validation cohort. Statistics for the enrichment analysis were evaluated with Student’s t-tests with ns=p>0.05, ***p ≤0.001, ****p ≤0.0001. **Abbreviations:** TB: tuberculosis; IGRA: Interferon-γ release assay; AUC: Area under curve; ROC: Receiver operating characteristic.

To investigate the association with commonly used clinical chemistry markers, we correlated the enrichment score of the 12-marker signature for TB disease, TB infection, and other non-TB disease separately with C-reactive protein (CRP), erythrocyte sedimentation rate (ESR), hemoglobin (Hb), albumin (Alb), and white blood cell count (WBC). In TB disease, both CRP and ESR correlated highly with the enrichment score (r = 0.69, and 0.65, respectively). Hb and Alb were negatively correlated (r = –0.47, and –0.79, respectively), while WBC was not correlated to the signature enrichment score (**Supplementary Figure 2**). Meanwhile, these correlations were lower in non-TB disease and TB infection across different markers.

As a next step, we trained a supervised prediction model using the signature profile of the discovery cohort and then tested it on the validation cohorts (**Figure 2, bottom panel**). The receiver operating characteristic (ROC) curves show the sensitivity and specificity of the signature in detecting individuals with TB disease versus TB infection. The target test performance, as indicated by the WHO TPP for a TB triage test, is defined by a minimum sensitivity of 90% and a minimum specificity of 70%, illustrated by the green box in each ROC analysis plot. The signature achieved a sensitivity of 95% (at a specificity of 70%) for TB disease in the discovery cohort (AUC = 0.97, CI: 0.92-1) (**Figure 2A**), indicating that the model is well-trained. However, as expected from the ssGSEA results, sensitivity was lower at 82% in the combined validation cohort (AUC = 0.82, CI: 0.75-0.9) (**Figure 2B**). Sub-group analysis showed superior accuracy in the Italian validation cohort (sensitivity 94%, AUC = 0.92, CI:0.86-0.97) compared with what was observed in the Swedish validation cohort (sensitivity 73%, AUC = 0.77, CI: 0.64-0.89) (**Figure 2C-D**).

### Optimization and reduction of the signature for enhanced accuracy

To further optimize and condense the 12-marker signature for easier clinical implementation, we used the Boruta feature selection algorithm in the discovery cohort to identify potentially redundant proteins. We found that 5 out of 12 proteins had low importance levels (**Supplementary** Figure 3) and were therefore removed. Among the remaining 7 proteins, CCL19 had a distinctly lower importance level compared to the other 6 proteins, enabling us to select and further validate a 6-marker signature: IL6, CDCP1, VEGFA, MCP-3, IFN-γ and CXCL9. The 6-marker signature was enriched in TB disease versus TB infection (**Figure 3, top panel**), with a higher performance in the validation cohorts (AUC=0.85, CI: 0.79-0.91) compared to the original 12-protein signature (AUC=0.82, CI: 0.75-0.9). Likewise, the 7-marker signature (6-marker signature plus CCL19) had a comparable performance (**Supplementary** Figure 4).

**Figure 3:**
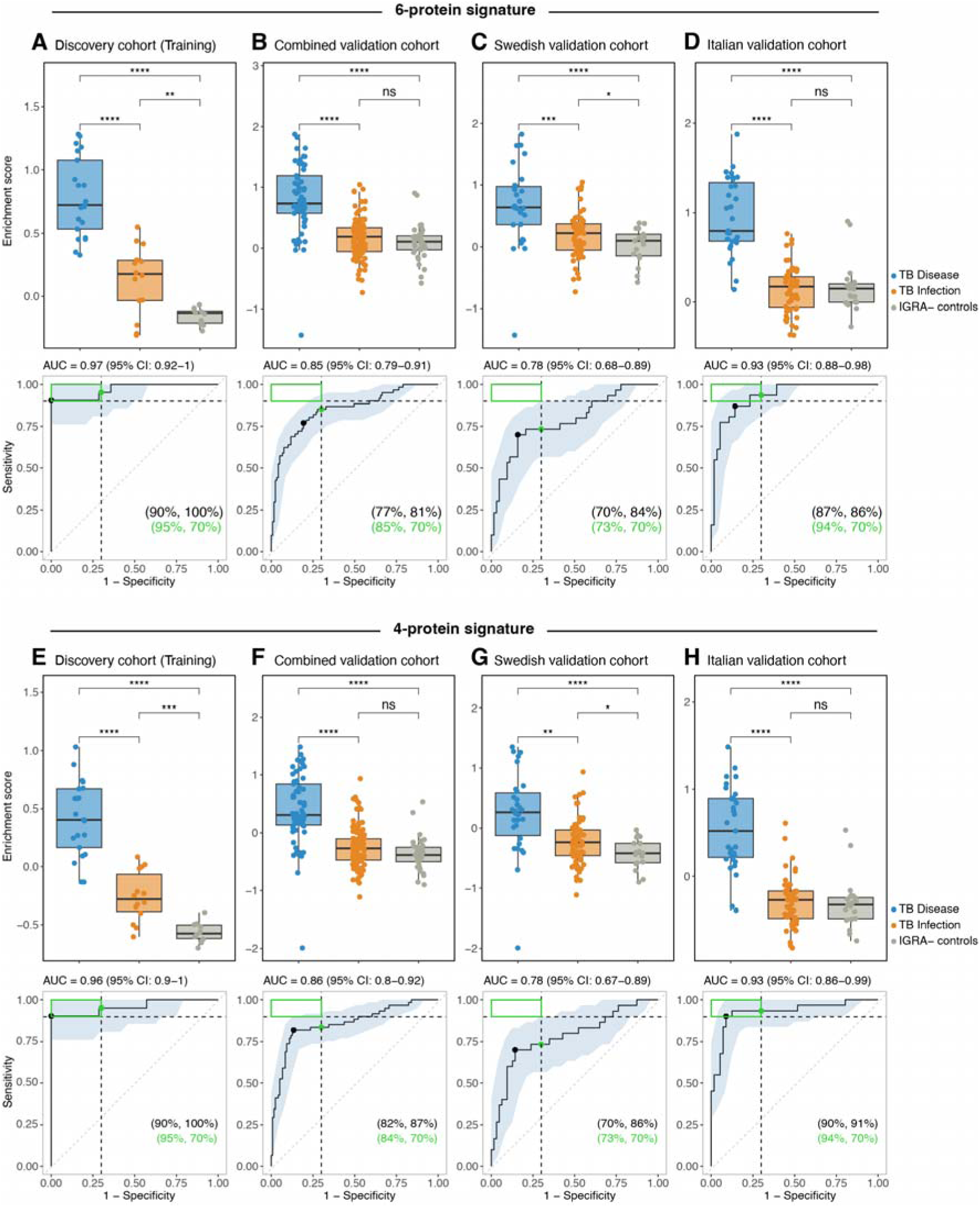
Accuracy of 6-protein and 4-protein signatures for diagnosis of TB disease. The first panel (**A–D**) represents the 6-protein signature, while the second panel (**E–H**) represents the 4-protein signature. The top row in each panel indicates single sample gene set enrichment analysis while the bottom row indicates ROC analysis, which was performed to classify TB disease versus TB infection. The shaded area indicates 95% confidence interval of the ROC curve while the green box in the upper left corner indicates the minimum requirement for a triage test (sensitivity: 90**%,** specificity: 70%). The sensitivity at a fixed specificity of 70% for TB disease detection is indicated by a green point with percent in the bottom right corner of the graph. The black point indicates the Youden index. The area under the ROC curve (AUC) with 95% confidence interval is indicated above each graph. (**A, E**) Training cohort, (**B, F**) combined validation cohort, (**C, G**) Swedish validation cohort, and (**D, H**) Italian validation cohort. Statistics for the enrichment analysis were calculated using a Student’s t-test with ns = p>0.05, *p≤0.05, **p≤0.01, **p≤0.001, ****p≤0.0001. **Abbreviations:** TB: tuberculosis; IGRA: Interferon-γ release assay; AUC: Area under curve; ROC: Receiver operating characteristic.

Next, we used the LASSO algorithm as another feature selection method to further optimize the 6-marker signature. According to the LASSO results (**Supplementary** Figure 5), a combination of four proteins had the lowest binomial deviance, minimizing the number of misclassified cases. We next proceeded to evaluate all 15 possible 4-marker combinations by performing ROC analysis (**Supplementary** Figure 6). We identified that the combination of IL6, CDCP1, IFN-γ, and CXCL9 had the highest accuracy in the validation cohort (AUC=0.86, CI: 0.80-0.92) (**Figure 3, bottom panel**). In addition to the 12-marker signature previously identified (10) and the 6-marker signature optimized by the Boruta algorithm, this 4-marker signature was also selected for further evaluation in the current study. As shown in **Figure 3**, the 4-marker and the 6-marker signature had comparable sensitivity (84% versus 85%) with a specificity fixed at 70%. Moreover, both signatures were significantly enriched in individuals with TB disease compared to the other groups, while no difference was observed between the TB infection and IGRA-negative controls.

### Signature enrichment in the non-TBD validation cohort

An important clinical question was whether these signatures were also enriched when comparing TB disease to non-TB disease (non-TBD). Both validation cohorts included individuals with non-TBD, TB infection, and TB disease, enabling us to assess the signature enrichment in the different groups. However, since the training dataset did not include non-TBD samples, hindering its utility in a train and test analysis, we used half of the samples from the combined Swedish and Italian validation cohorts as a training set and the other half for validation.

The three signatures were significantly enriched in TB disease compared to non-TBD, with little difference observed between TB infection and non-TBD (**Figure 4A-C**). ROC analysis demonstrated a performance (AUC) ranging between 0.76-0.79 for the 12-marker, 6-marker, and 4-marker signatures in distinguishing TB disease from non-TBD individuals. The 4-marker signature showed the highest AUC (AUC=0.79; 95% CI: 0.66-0.91) in this comparison and achieved a sensitivity of 81% at a specificity of 70%. To further examine the relative abundance of the 12 proteins within each group, we generated a heatmap showing the Z-score of the abundance level of each protein across individual donors in different groups (**Figure 4D**).

**Figure 4:**
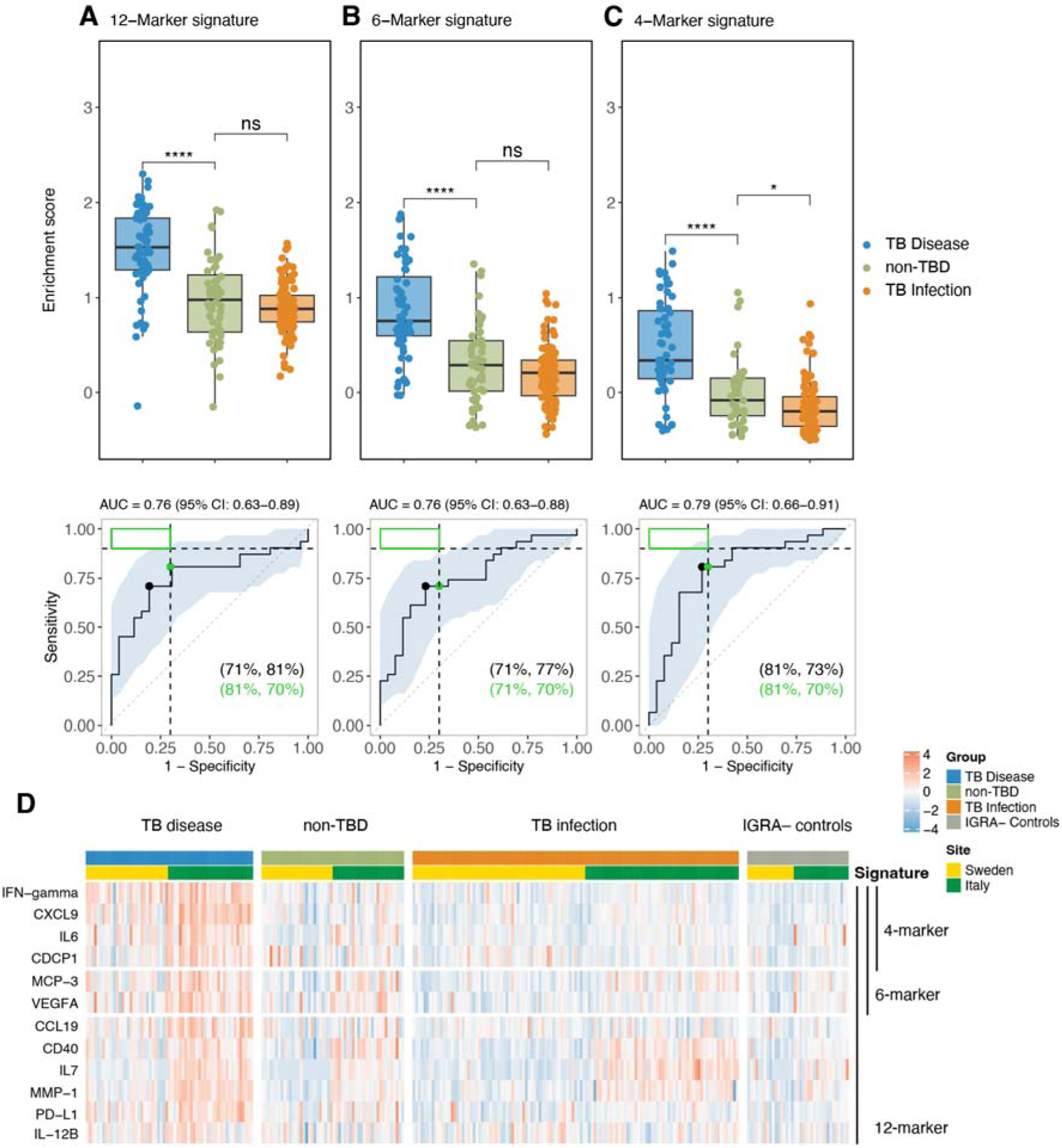
Signature enrichment in TB disease compared with other relevant non-TB diseases. Single sample gene set enrichment analysis (top panel) for the 12-marker (**A**), 6-marker (**B**) and 4-marker (**C**) signature in TB disease versus other relevant non-TB disease (non-TBD), and TB infection. The bottom panel represents ROC analysis of the different signatures classifying TB disease versus non-TBD. (**D**) Heatmap showing Z-score normalized relative protein abundance levels for each donor, ordered by group (X-axis) and signature (Y-axis), where red indicates higher relative abundance and blue indicates lower relative abundance within each row. Statistics for the enrichment analysis were calculated using a Student’s t-test with ns = p>0.05, *p≤0.05, ****p≤0.0001. **Abbreviations:** non-TBD: non-TB Disease; TB: tuberculosis; IGRA: Interferon-γ release assay.

Since the anticoagulant can affect plasma protein levels, we extended the analysis to compare signature enrichment in both heparin and EDTA plasma in a subset of samples from the Swedish cohort (n=73, TB disease n=25, non-TBD n=25, TB infection n=23). All three signatures performed comparably, and the enrichment scores were highly correlated between heparin and EDTA (**Supplementary** Figure 7), although they varied at the individual marker level (**Supplementary** Figure 8), with IFNg (r=0.98), IL-6 (r=0.96), CXCL9 (r=0.93), IL-12B (r=0.93), and CDCP1 (r=0.9) having the highest correlation between heparin and EDTA. Comparing relative protein quantity (NPX values), all proteins except CXCL9, MCP-3, CD40, IL-7, and IL-12B, were present at comparable levels in both sample types (**Supplementary** Figure 9).

### Comparison with other published TB disease signatures

We have previously reviewed (9) published protein signatures associated with TB disease. The Olink Target 96 inflammation panel used in this study, containing 92 proteins, allowed us to compare ten of these signatures (11–21) with our own 12-, 6-, and 4-marker signatures for distinguishing TB disease from TB infection (**Figure 5**). First, we trained a supervised prediction model using the markers included in each signature on the training dataset (**Figure 5A**). We then evaluated the accuracy of each model in identifying TB disease from TB infection on the combined validation cohort and the Swedish and Italian validation cohorts separately (**Figures 5B-D**). Three signatures out of the ten, consisting of IFN-γ, ADA, and CXCL10 (13) (AUC=0.84), IFN-γ in combination with ADA (11) (AUC=0.84) and IFN-γ alone (11) (AUC=0.86) performed similarly to our 6- and 4-marker signatures (AUC=0.85 and 0.86, respectively).

**Figure 5:**
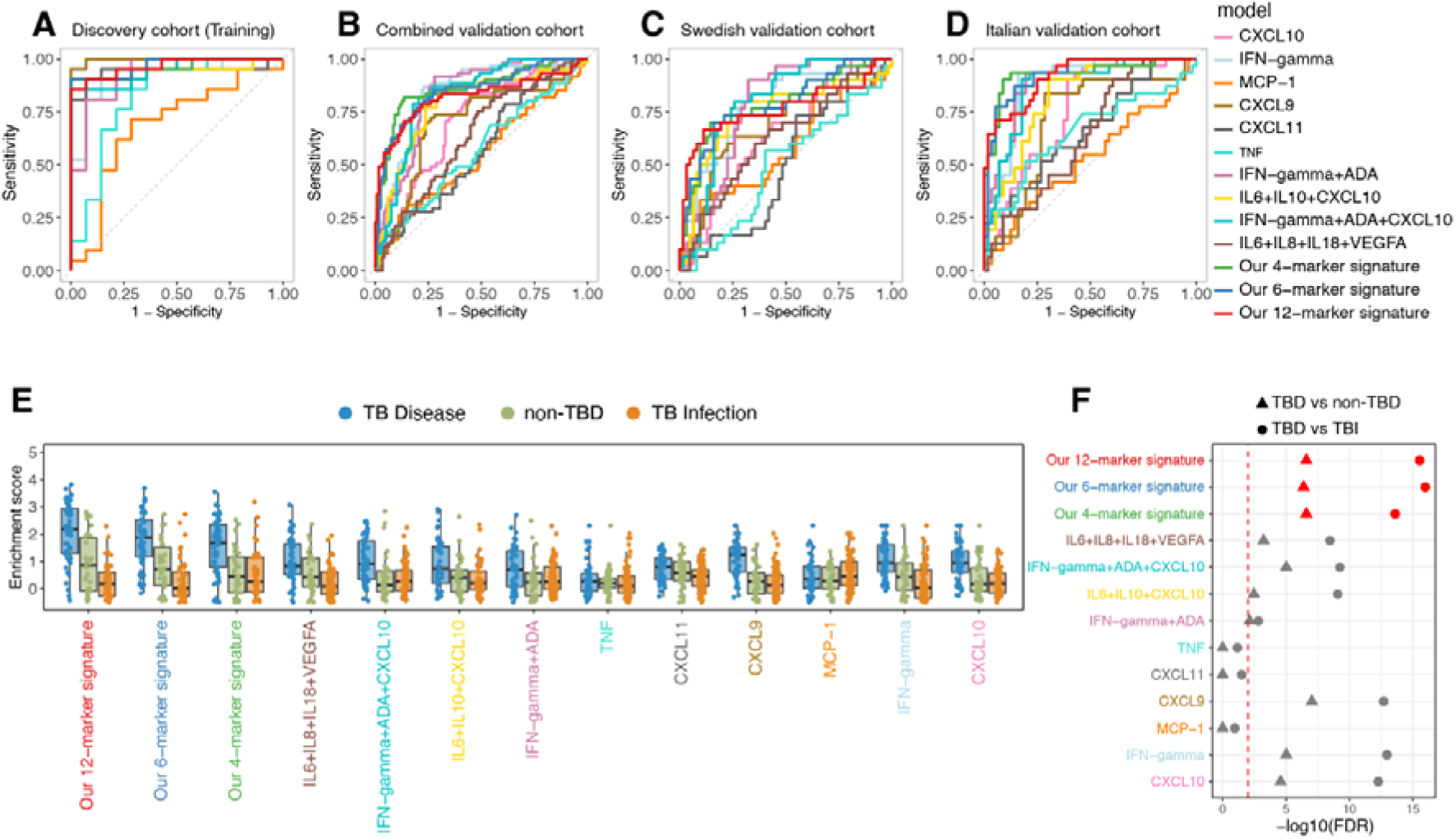
Comparison with other published TB disease signatures. Comparison of the 12-marker, 6-marker, and 4-marker signatures with 10 other published signatures/markers. (**A–D**) ROC analyses in different cohorts, with signatures/markers indicated by different colors for classifying TB disease versus TB infection. (**E**) Single sample gene set enrichment analysis results for the combined validation cohort, comparing TB disease with TB infection, and non-TBD. (**F**) Statistics for the enrichment analysis were calculated using Student’s t-test between TB disease and TB infection (circle) or TB disease and non-TBD (triangle) and corrected using FDR. The red dashed line indicates the FDR-corrected p = 0.01. **Abbreviations:** non-TBD: non-TB Disease; TB: Tuberculosis; IGRA: Interferon-γ release assay; ROC: Receiver operating characteristic; FDR: False Discovery Rate.

In addition to the ROC analysis, we calculated the enrichment score for each signature in individual samples to determine which signatures were enriched in TB disease versus non-TBD, or TB infection (**Figure 5E**). Ten of the evaluated signatures including IFN-γ+ADA+CXCL10, IFN-γ+ADA and IFN-γ alone were significantly enriched (FDR-corrected p < 0.01) in TB disease compared to TB infection, including our three signatures which demonstrated higher significance levels (**Figure 5F**). Comparing the enrichment score for each signature in TB disease vs. non-TBD, all ten signatures including our three signatures, remained significantly enriched (FDR-corrected p < 0.01) (**Figure 5F**).

Finally, we performed ROC analysis for each individual marker distinguishing TB disease from TB infection (**Supplementary** Figure 10), and from non-TB disease (**Supplementary Figure 11)**. Although when compared to the 4-protein signature, IFN-γ demonstrated slightly higher accuracy (sensitivity 89% vs. 84% at 70% specificity) in predicting TB disease vs. TB infection, its accuracy was much lower (sensitivity 58% vs. 81% at 70% specificity) in distinguishing TB disease vs. non-TB disease. Together, these findings indicate that multivariate host-response signatures are necessary to increase both sensitivity and specificity across clinically heterogeneous presentations of TB disease.

### High correlation between suspension bead array and proximity-extension assay methodologies for assessing plasma cytokine levels

The use of proximity extension assay (PEA)-based protein quantification has expanded greatly in recent years, with large datasets having been generated for a wide variety of diseases (22, 23). However, the technology primarily generates relative protein abundance levels and has been described as having a very sensitive protein detection, making it difficult to know if the levels detected are quantifiable using other more accessible platforms, such as suspension bead arrays. We therefore assessed CDCP1, CXCL9, IFN-γ, and IL6 levels for a subset of heparin samples using the EYRA (all proteins), Luminex (IFN-γ and CXCL9) and Legendplex (IL6) platforms. Arrays including CDCP1 were not found commercially, so an in-house assay was set up. All four proteins were significantly higher in individuals with TB disease compared with TB infection and for three of the proteins (CXCL9, IFN-gamma, and IL6) also against non-TBD (**Figure 6A**). This was the case for all bead platforms (**Supplementary** Figure 12). The plasma protein levels between the assays were well correlated for CXCL9 (r=0.75), IFN-γ (r=0.63), and IL6 (r=0.81), but less for CDCP1 (r=0.25). The reduced correlation for CDCP1 was primarily due to plasma levels falling below the lower limit of detection for the assay (study. As shown in and **Supplementary Figure 12**).

**Figure 6:**
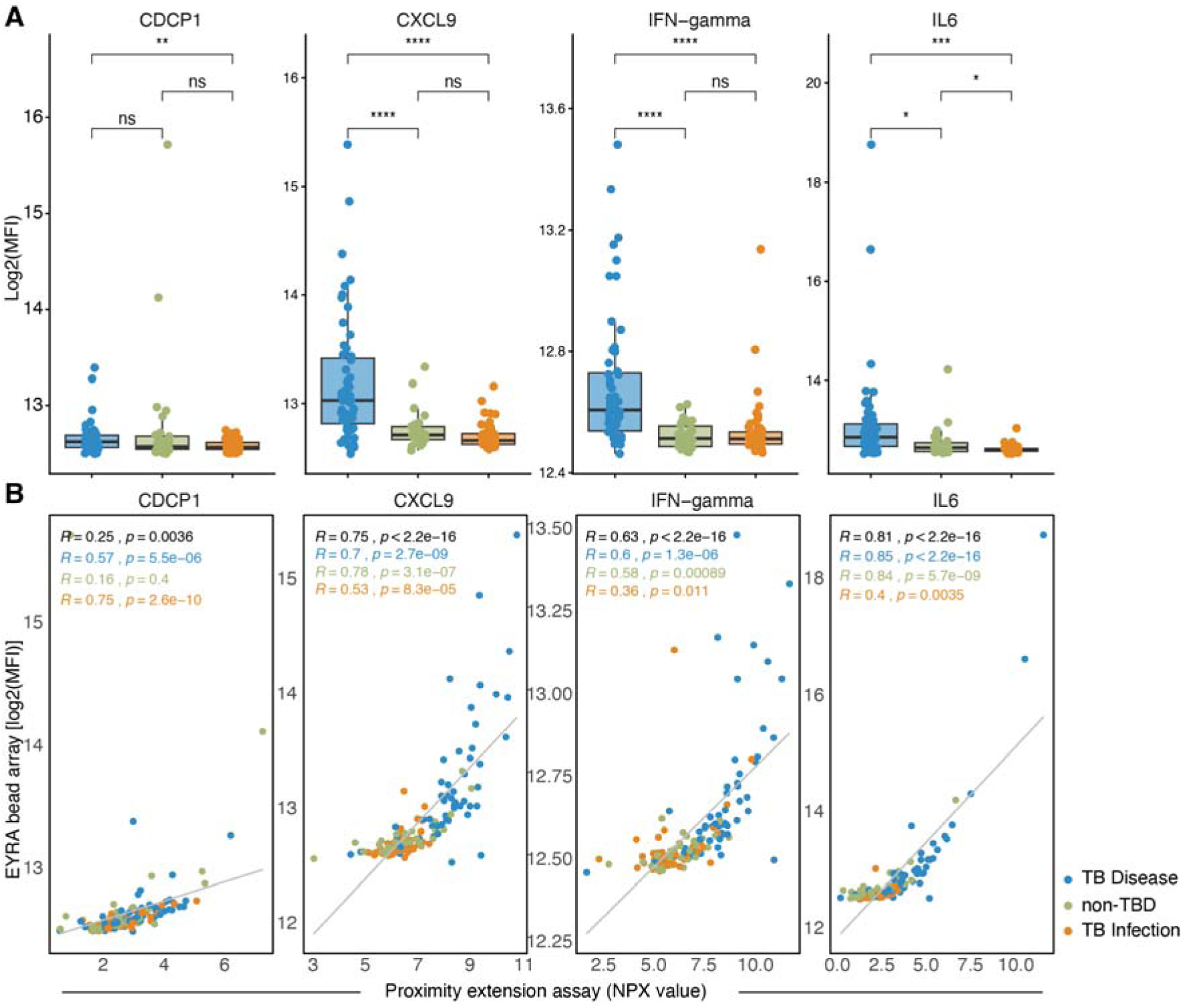
Cytokine levels quantified by suspension bead array and correlation with proximity-extension assay. (**A**) CDCP1, IFN-gamma, CXCL9, and IL6 levels were quantified by suspension bead array (EYRA) in heparin plasma from a subset of donors with TB disease (blue, n=59), non-TBD (green, n=36), and TB infection (orange, n=57). Cytokine levels were compared between groups using the Student’s t-tests with ns = p>0.05, *p<0.05, **p<0.01, ***p<0.001, ****p<0.0001. (**B**) Pearson correlation (r) was calculated between Log2 transformed suspension bead array-derived cytokine MFI (median fluorescent intensities) and proximity extension assay (Olink) normalized protein expression (NPX) values.

## Discussion

We conducted an external validation of a previously described 12-marker host-response protein signature related to TB disease severity (10). In addition, we condensed and optimized the signature into a 6- and a 4-marker signature, both showing comparable, or modestly improved, performance relative to the original signature.

We used two validation cohorts, one Swedish and one Italian. The sensitivity of the 12-marker signature was higher in the Italian cohort (94% at 70% specificity) than in the Swedish cohort (73%). We propose that this difference is likely influenced by the broader range of TB disease severity in the Swedish cohort consisting of a mixture of pulmonary and extrapulmonary TB. Many Swedish individuals did not report any general symptoms (40%; 12 of 30). Moreover, 47% of those with pulmonary TB disease required invasive investigations, such as bronchoscopy, to verify the diagnosis, indicating a lower bacterial burden. Four individuals were identified via contact tracing and were thus diagnosed early during their TB disease, with very mild clinical symptoms. All four displayed low signature enrichment and inflammatory biochemistry markers. This contrasted with the Italian TB cohort, consisting of individuals with pulmonary TB with a higher smear-positivity rate (81% vs 41% in the Swedish cohort) and an overall higher smear-grade, with extensive lung lesions (as evaluated by chest-X ray) indicating more extensive disease.

This pattern is consistent with our previous observation that the 12-marker signature captures a TB-associated inflammatory state linked to disease severity (10). Thus, a stronger signature was associated with biochemical markers indicating more extensive inflammation, such as lower Hb and albumin, and higher ESR and CRP. The presence of any systemic symptoms (fever, night sweats, loss of appetite and weight loss) and smear positivity was also associated with a stronger signature (10). This aligns with prior evidence that pulmonary TB is characterized by increased circulating proinflammatory cytokines correlating with bacterial burden and disease extent (24).

The condensed 4-marker (IL6, CDCP1, IFN-γ and CXCL9) and 6-marker (IL6, CDCP1, VEGFA, MCP-3, IFN-γ and CXCL9) signatures had a similar accuracy with a sensitivity of 84% (AUC=0.86) and 85% (AUC=0.85), respectively at a specificity of 70% overall. While the 12-marker signature reached an overall sensitivity of 82% (AUC=0.82) at the same specificity. In the Italian validation cohort, all three identified signatures reached the WHO TPP 90% sensitivity triage criteria: 94% (12-marker, AUC=0.92), 94% (6-marker, AUC=0.93) and 94% (4-marker, AUC=0.93) sensitivity at 70% specificity. These findings suggest that the signatures may be informative in the context of pulmonary TB, particularly in settings where patients present with more advanced inflammatory disease, although this observation warrants further targeted validation. Encouragingly, the 12-protein signature demonstrated the same enrichment score in extrapulmonary TB, a condition in which blood-based biomarkers would be particularly advantageous given the reliance on invasive diagnostic procedures.

The 2014, WHO TPPs for TB (25) include both a screening and a diagnostic test for TB disease, where we have matched our results against the former using the minimum requirement of 90% sensitivity and 70% specificity for confirmed pulmonary TB. In 2024, an updated TPP (26) was released for diagnosis of TB disease intended for use at peripheral health clinic level in TB endemic areas. Performance metrics in relation to complexity of the analysis were emphasized and programmatic considerations, such as the ease of handling, turn-around time, and cost per test were introduced as important factors in the diagnostic guidelines. So far, blood-based assays built on transcriptomic expression (e.g. Xpert MTB-HR) have nearly reached the TPP performance specifications for a screening test (27, 28), but the operational requirements are yet to be reached. An assay for protein-detection has several advantages as there are platforms such as lateral flow assays (29) that enable point-of-care use at lower costs. The stepwise reduction of the signature from 12 to 4 proteins represents an initial proof-of-concept toward compatibility with simpler analytical formats, rather than readiness for clinical implementation.

Given that the primary clinical application of blood-based tests for TB disease is in symptomatic individuals, among whom TB represents one of several differential diagnoses, we evaluated whether these signatures were also enriched in non-TB diseases associated with overlapping immune responses. All three signatures (12-, 6-, and 4-marker) showed significantly greater enrichment in TB disease than in non-TB diseases. These findings are consistent with our previous report (10), in which the 12-marker signature was likewise significantly enriched in TB disease relative to non-TB diseases across published transcriptional datasets (10). The sensitivity of the 4-marker signature was 81% at a specificity of 70% with an AUC of 0.79 in the combined validation cohort. This accuracy is comparable to that reported for transcriptomic host-response assays such as Xpert MTB-HR in similar clinical settings (30), further indicating an overlap between the protein-based signatures and gene expression signatures for TB disease. In the Folkesson et al. study, assay performance was also strongly associated with sputum bacterial load and clinical presentation (30).

In addition to the signatures described here, a small number of other promising protein-based TB signatures have been validated in independent cohorts. Among those meeting the WHO target product profile criteria for a blood-based diagnostic or triage test for TB disease, only three have been validated, with variable results (9). Two were evaluated in serum samples (31,32), and one in plasma (33). The signature of Chegou et al. (31) did not reach the TPP criteria upon validation, with a sensitivity of 89% and a specificity of 60% in serum (32) while the signature of De Groote et al. (33) was validated in a blinded set of serum samples provided by FIND, with a sensitivity of 85% and specificity of 89% (33). The signature of Zhao et al. (34) was validated in two independent test sets in plasma; In test set A the signature was successfully validated, with no significant change in the AUC (0.95) compared to the training set (1.0) while in test set B, the AUC was reduced to 0.78 (34).

An additional aspect is to assess signature performance in different sample types. Samples collected with different anti-coagulants have previously been shown to affect protein levels (35–38). In this study, we compared the signature enrichment in heparin and EDTA plasma. Importantly, all three signatures performed comparably, and the enrichment scores were highly correlated between heparin and EDTA. Of the 12 proteins included in the original signature, 3 proteins (CXCL9, MCP-3, CD40) were higher in plasma with heparin while 2 proteins (IL-7, IL-12B) were higher in EDTA. The variations seen were relatively minor for CXCL9 and CD40, while the others had levels over two-fold change. There are few other studies with overlapping analytes that assess the effect of anticoagulants. In a study of Patil et al. (36) IL-6 was included, and in contrast to here, the level was significantly higher in plasma with heparin than in plasma with EDTA. Biancotto et al (38) found that MCP-1, a CC chemokine with similar activity as chemoattractant MCP-3 in our study, also showed significantly higher concentrations in heparinized plasma compared to EDTA plasma. Importantly though, the differences for individual proteins did not affect the high correlation seen between heparin and EDTA samples for the signature itself.

In conclusion, the 12-marker TB disease signature was reduced to 6- and 4-marker formats without compromising accuracy. Both optimized signatures also showed significant enrichment in TB disease relative to non-TB disease. Across multiple comparative analyses against ten published signatures, the condensed 6- and 4-marker signatures consistently achieved high diagnostic accuracy. A major strength of this study is the validation of these signatures in independent cohorts encompassing a broad spectrum of disease severity and co-morbidities. The signatures were also benchmarked against published signatures and assessed across different sample types and assay platforms within the same patient cohorts. The main limitations include the relatively small sample size and the limited number of individuals with non-TB disease presenting with TB-like symptoms. Nonetheless, the consistent findings in both pulmonary and extrapulmonary TB support further validation of the 6- and 4-marker signatures in larger cohorts of individuals with presumed TB across diverse clinical settings, particularly in TB-endemic areas.

## Methods

### Clinical cohorts

#### Ethical considerations

All study participants received written and verbal information and signed a written informed consent form prior to study inclusion. In relevant cases professional interpreter services were used.

The Swedish study cohort received ethical permission from the Swedish Ethical Review Authority, EPN-number 2019-05438 with addendums 2022-00231-02, 2022-07053-02 with ÖN 29-2023, and 2024-03955-02. The Italian study cohort was approved by the Ethical Committee of the National Institute for Infectious Diseases Lazzaro Spallanzani-IRCCS (approval number 72/2015 and amendment approval number 27/ 2019). After obtaining the clinical and demographic information, the samples from Italy were anonymized.

#### Swedish study cohort

Adults (>18 years old) with TB disease or TB infection were prospectively recruited at convenience at the Infectious Disease Department, Karolinska University Hospital, Stockholm between May 2018 and November 2023. Inclusion criteria: 1) Individuals with presumed or confirmed TB disease with blood samples taken within 1 week of anti-TB treatment initiation. TB disease was microbiologically verified via *Mtb* culture or *Mtb* PCR in sputum and non-sputum samples. 2) Individuals with TB infection defined as a positive IGRA result (QuantiFERON-TB Gold In-Tube [QFT-GIT] or QuantiFERON-TB Gold Plus [QFT-Plus]) mainly falling into three categories: i) close contacts of an index patient with infectious pulmonary TB; ii) screening of migrants from a TB high-endemic country: (iii) screening of patients in anticipation of immunosuppression or in the peripartum period. 3) Individuals with clinically relevant non-TB disease (non-TBD); presenting to the clinic as presumed TB disease but in whom TB disease was ruled out by chest X-ray, medical examination and mycobacterial sampling. 4) IGRA-negative controls consisted of individuals investigated for TB infection who upon follow-up had negative IGRA results (**Supplementary Tables 1 and 2**).

#### Italian study cohort

Patients and healthy donors were prospectively enrolled from 2016 to 2022. TB infection diagnosis was determined based on a positive QuantiFERON (QFT)-Plus assay result (Diasorin, Vercelli, Italy), without clinical, microbiological, or radiological evidence of TB disease. They were enrolled before starting preventive treatment. TB disease patients had a microbiological diagnosis (either by culture or microbiological means) and were enrolled either before starting treatment or within 7 days of therapy initiation. As controls, healthy individuals who tested negative for QFT-Plus were enrolled. Non-TB disease included other respiratory disease patients who were initially suspected of having TB but were later diagnosed with a different condition after comprehensive clinical, microbiological, and radiological evaluations.

The demographic and clinical characteristics of all cohorts used in this study are reported in **Supplementary Table 3**. Due to the observational nature of this study with unpredictable outcomes, a convenient sample of subjects was selected, considering laboratory workflow, enrollment duration, patient flow in the hospital, and experimental protocol costs.

#### Sample preparation

Venous blood was collected into heparin or EDTA vacutainer tubes and transferred to the laboratory for immediate preparation. The tubes were centrifuged at 670 *g* for 8 min (Swedish cohort) or at 409 *g* for 10 min (Italian cohort). The plasma fraction was then aliquoted and stored at –80°C.

#### CDCP1, IFN-gamma, CXCL9, and IL6 quantification by EYRA suspension bead array

CDCP1, IFN-gamma, CXCL9, and IL6 levels were determined using a multiplex EYRA assay (Mabtech) according to the manufacturer’s instructions. Briefly, beads were washed four times with the provided wash buffer. Then heparin or EDTA plasma samples, diluted 1:2 in Assay diluent, were added and incubated with the beads for 2 h on an orbital shaker set at 800 rpm. The beads were then washed four times with wash buffer, followed by a 1 h incubation with the detection antibody mix on an orbital shaker set at 800 rpm. After washing the beads four times, they were incubated for 30 min with Streptavidin-PE on an orbital shaker set at 800 rpm. Beads were washed four times and resuspended in 50 µl of Assay diluent by pipetting and by incubation on an orbital shaker set at 800 rpm for 5 min. After letting the beads settle for 10 min, the plate was read on the EYRAplex machine (Opal2 build53) and concentrations calculated through interpolation from standard curves.

In addition, an in-house assay for CDCP1 was set up based on validated antibody pairs (Detection antibody clone EPR22487-51 and capture antibody clone EPR22487-289, both from Promega). The detection antibody was biotinylated using the Biotinylation Kit / Biotin Conjugation Kit (Fast, Type A) (Lightning-Link) according to the manufacturer’s instructions. The capture antibody was conjugated to EYRA beads with an adapted protocol from Mabtech. Beads were washed four times and resuspended in milliQ water before adding S-NHS (Thermo Fisher) and EDAC (Sigma Aldrich), both at 50 mg/ml in milliQ water. The reaction tube was then incubated for 15 min on an oscillating rotator at 25 rpm at RT. The beads were washed four times and resuspended in conjugation buffer (milliQ water at pH 5 set with HCl and NaOH), prior to adding the capture antibody in a ratio of 3.75 µg/million beads. The reaction was incubated o/n at RT on an oscillating rotator at 25 rpm. On day two, the beads were washed once with conjugation buffer, followed by three times with quenching buffer (1xPBS with 100 mM ethanolamine, pH 8 set with HCl and NaOH) and resuspended in quenching buffer before incubation for 1h at RT on an oscillating rotator at 25 rpm at RT. After washing and resuspending the beads in storage buffer (1xPBS with 1% BSA, pH 7.4), the bead concentration was determined using an automated cell counter. The reaction tube was protected from light during all incubations.

#### IFN-gamma and CXCL9 quantification by Luminex suspension bead array

IFN-gamma and CXCL9 concentrations were determined using a custom multiplex Luminex assay (R&D Systems) according to the manufacturer’s instructions. Briefly, heparin plasma was diluted in Calibrator Diluent RD6-52 buffer followed by incubation with beads for 2 h on an orbital shaker set at 800 rpm. The beads were then washed three times with provided wash buffer, after which they were incubated with biotinylated cytokine specific detection antibodies for 1 h on the shaker. After washing three times with wash buffer, beads were incubated with streptavidin-PE for 30 min. After a final set of washes, the beads were read on a Bioplex-200 machine and concentrations calculated through interpolation from a standard curve.

#### IL-6 quantification by LegendPlex suspension bead array

IL-6 was quantified using the multi-analyte flow assay kit for LegendPlex (BioLegend 50-208-8603) in a v-bottom 96 well plate according to the manufacturer’s instructions. Briefly, samples were diluted in provided assay diluent immediately preceding incubation with magnetic beads for 2 h on an orbital shaker set at 800 rpm. Beads were washed twice with provided wash buffer before the addition of detection antibodies and 1 hour incubation on an orbital shaker set at 800 rpm. Beads were washed twice again, followed by incubation with streptavidin-PE for 30 min on an orbital shaker set at 800 rpm. Beads were washed twice and resuspended in 150 µL of wash buffer before reading on a 3 Laser Cytek Aurora flow cytometer at low acquisition speed. IL-6 concentrations were determined from fcs files using interpolation from a standard curve provided by Legendplex Qognit software.

### Analysis of proteins with Olink PEA technology

The Proximity Extension Assay (PEA) provided by Olink was used for protein profiling (39). In this technology, two paired antibodies, each conjugated with a matched oligonucleotide, target the same protein. When the antibodies are in close proximity, the matched oligonucleotides hybridize, forming a template that serves as a primer for DNA polymerase. This is then extended and amplified using either Polymerase Chain Reaction (PCR) or Next Generation Sequencing (NGS) technology. Using this technology, we profiled the abundance of 92 inflammation-associated protein in plasma using the Olink Target 96 Inflammation panel (Olink).

### Quantification and statistical analysis

#### Protein abundance data analysis (Olink preprocessing)

For each protein, the NPX value, which is a log2-transformed abundance level, was used initially. Since the samples in this study were run across different batches, we included some overlapping samples in each dataset to enable bridge normalization and prevent batch effects in the final dataset. We used the read_NPX and olink_normalization_bridge functions from the OlinkAnalyze R package to import the data and perform bridge normalization.

#### Feature selection algorithms (Boruta and LASSO)

For feature optimization, we first used the Boruta algorithm (40) followed by LASSO (41) sequentially. Boruta is a random forest-based feature selection algorithm that identifies features significantly contributing to classification. LASSO (Least Absolute Shrinkage and Selection Operator) is a supervised learning approach that performs both regularization and feature selection, allowing us to identify the most relevant features by shrinking less important ones to zero. The sequential combination of these two algorithms enhances the robustness of the selected feature set by first narrowing down important features with Boruta, followed by further refinement using LASSO. We used the Boruta and glmnet R packages to implement Boruta and LASSO, respectively.

#### Gene set enrichment analysis (ssGSEA)

We used the ssGSEA (single-sample Gene Set Enrichment Analysis) method (42) to assess the activity of markers from each signature against the entire set of proteins and compare them between different groups of samples. GSEA performs a statistical test to indicate how significantly a set of genes is enriched in one group versus another. However, with ssGSEA, we can obtain the enrichment score for each individual sample rather than for group comparisons. We used the corto R package to run ssGSEA. To compare enrichment scores between groups, we used Student’s t-test to detect significant differences in mean enrichment scores.

#### ROC analysis (support vector machine)

To assess the accuracy of each signature, we trained an SVM (Support Vector Machine) model using the training dataset and then evaluated the model on the test dataset. SVMs are particularly advantageous for proteomic data, where nonlinear relationships among proteins may be present. The ROC (Receiver Operating Characteristic) curve was used to demonstrate the accuracy of each model in terms of sensitivity and specificity across different thresholds. In the ROC curve, we selected a specific threshold aiming for a minimum sensitivity of 90% while specificity was set to 70%, in accordance with the WHO TPP (Target Product Profile) criteria for a TB triage test (25). We used the caret and pROC R packages to implement SVM and ROC analysis, respectively.

## Acknowledgments

We would first like to thank all study participants. We would also like to express our gratitude to all clinicians, nurses, laboratory staff, and students for assisting us with study inclusion, sampling, and processing. We are thankful to the Affinity Proteomics Unit at SciLifeLab Stockholm for the generated Olink data.

This study was supported by grants from the Swedish Research Council (2021-03706 and 2023-01943), the Swedish Medical Association (SLS-934363 and SLS-960484), the Swedish Society for Medical Research (CG-24-0012-B), and the Heart-Lung Foundation (20220566, 20230244, and 20250418) to CS. Grants from Swedish Research Council (2019-04663 and 2020-03602) and the Heart-Lung Foundation (20180386 and 20200194) to GK. ZM was supported by grants from the VALIDATE Network and the Erik and Edith Fernström Foundation for Medical Research. The Italian team was funded by the Italian Ministry of Health (Ricerca Corrente, Linea 4, Progetto 2), and TBVACHORIZON, funded by the European Union’s HORIZON program under Grant No. 101080309.

## Author contributions

ZM, CS, and GK designed the study. EF and JB included patients and collected clinical information for the Swedish cohort. XZ, LK, and CSS prepared the Swedish samples. XZ, MSG, LB, and LK performed assays. VV prepared the Italian samples; GC managed the Italian database. DG, GG and FP enrolled the subjects from the Italian cohort. ZM performed data analysis. ZM, EF, GC, DG, and CS generated figures and tables. ZM, EF, DG, JB, GK, CS interpreted the results. CS, GK, JB, ZM and DG provided funding. CS and GK provided project supervision and mentoring. ZM, EF, CS, and GK wrote the initial draft with input from DG, MCN, and JB. All authors contributed to revision and editing.

## Data availability statement

Data is available upon reasonable request to the corresponding author and pending confirmation of relevant permits to access GDPR protected pseudoanonymized data. Analysis code is available via GitHub: https://github.com/SundlingLab/Olink_ATBsignature_Validation.

## Supplemental information

**Supplementary Table 1**: Demographic and clinical information of study participants from Sweden

**Supplementary Table 2:** Clinical characteristics of Swedish study participants with TB disease

**Supplementary Table 3:** Demographic and clinical information of study participants from Italy

**Supplementary Figure 1:** Enrichment score of the 12-marker signature in pulmonary TB and extrapulmonary TB and early and minimal symptomatic disease.

**Supplementary Figure 2.** Correlation between enrichment scores from the 12-marker signature and various clinical blood chemistry markers.

**Supplementary Figure 3.** The importance level of each marker from the 12-protein signature in classifying TB disease versus TB infection, as calculated by Boruta.

**Supplementary Figure 4.** Accuracy of the 7-marker signature selected based on the Boruta output, for TB disease.

**Supplementary Figure 5.** Binomial deviance for each combination of the 6-marker signature selected by Boruta, calculated using LASSO regression.

**Supplementary Figure 6.** ROC plot for all combinations of 4 markers out of 6 markers, selected based on the Boruta output.

**Supplementary Figure 7.** Anticoagulant effects on the performance of the 12-marker, 6-marker and 4-marker signatures.

**Supplementary Figure 8.** Pearson correlation between the enrichment score from heparin and EDTA samples for each protein included in the 12-marker signature.

**Supplementary Figure 9.** Comparison between NPX values for heparin and EDTA the individual proteins of the 12-marker signature.

**Supplementary Figure 10.** ROC analysis for each protein of the 12-marker signature for classifying TB disease versus TB infection.

**Supplementary Figure 11.** ROC analysis for each protein of the 12-marker signature for classifying TB disease versus non-TB disease.

**Supplementary Figure 12.** Suspension bead array results with concentrations for IFN-gamma, IL6, and CXCL9 based on Luminex and Legendplex systems.

## REFERENCES

1. WHO. Global Tuberculosis Report. https://www.who.int/publications/i/item/9789240101531. 20.

2. Churchyard G, Kim P, Shah NS, Rustomjee R, Gandhi N, Mathema B, et al. What We Know About Tuberculosis Transmission: An Overview. The Journal of infectious diseases. 2017;216(suppl_6):S629–S35.

3. Norbis L, Alagna R, Tortoli E, Codecasa LR, Migliori GB, Cirillo DM. Challenges and perspectives in the diagnosis of extrapulmonary tuberculosis. Expert Rev Anti Infect Ther. 2014;12(5):633–47.

4. Jain R, Gupta G, Mitra DK, Guleria R. Diagnosis of extra pulmonary tuberculosis: An update on novel diagnostic approaches. Respir Med. 2024;225:107601.

5. Goletti D, Al-Abri S, Migliori GB, Arlehamn CL, Haldar P, Sundling C, et al. World Tuberculosis Day 2024 theme “Yes! We can end TB” can be made a reality through concerted global efforts that advance detection, diagnosis, and treatment of tuberculosis infection and disease. International Journal of Infectious Diseases. 2024;141.

6. Organization WH. WHO consolidated guidelines on tuberculosis. Module 3: diagnosis–rapid diagnostics for tuberculosis detection: World Health Organization; 2024.

7. Kontsevaya I, Cabibbe AM, Cirillo DM, DiNardo AR, Frahm N, Gillespie SH, et al. Update on the diagnosis of tuberculosis. Clin Microbiol Infect. 2024;30(9):1115–22.

8. Nogueira BMF, Krishnan S, Barreto-Duarte B, Araujo-Pereira M, Queiroz ATL, Ellner JJ, et al. Diagnostic biomarkers for active tuberculosis: progress and challenges. EMBO Mol Med. 2022;14(12):e14088.

9. Mousavian Z, Kallenius G, Sundling C. From simple to complex: Protein-based biomarker discovery in tuberculosis. Eur J Immunol. 2023;53(12):e2350485.

10. Mousavian Z, Folkesson E, Froberg G, Foroogh F, Correia-Neves M, Bruchfeld J, et al. A protein signature associated with active tuberculosis identified by plasma profiling and network-based analysis. iScience. 2022;25(12):105652.

11. Kalantri Y, Hemvani N, Chitnis DS. Evaluation of real-time polymerase chain reaction, interferon-gamma, adenosine deaminase, and immunoglobulin A for the efficient diagnosis of pleural tuberculosis. Int J Infect Dis. 2011;15(4):e226–31.

12. Hong JY, Jung GS, Kim H, Kim YM, Lee HJ, Cho SN, et al. Efficacy of inducible protein 10 as a biomarker for the diagnosis of tuberculosis. Int J Infect Dis. 2012;16(12):e855–9.

13. Sutherland JS, Garba D, Fombah AE, Mendy-Gomez A, Mendy FS, Antonio M, et al. Highly accurate diagnosis of pleural tuberculosis by immunological analysis of the pleural effusion. PloS one. 2012;7(1):e30324.

14. Lee K, Chung W, Jung Y, Kim Y, Park J, Sheen S, et al. CXCR3 ligands as clinical markers for pulmonary tuberculosis. The international journal of tuberculosis and lung disease : the official journal of the International Union against Tuberculosis and Lung Disease. 2015;19(2):191–9.

15. Wergeland I, Pullar N, Assmus J, Ueland T, Tonby K, Feruglio S, et al. IP-10 differentiates between active and latent tuberculosis irrespective of HIV status and declines during therapy. The Journal of infection. 2015;70(4):381–91.

16. Santos AP, Correa RDS, Ribeiro-Alves M, Soares da Silva ACO, Mafort TT, Leung J, et al. Application of Venn’s diagram in the diagnosis of pleural tuberculosis using IFN-gamma, IP-10 and adenosine deaminase. PloS one. 2018;13(8):e0202481.

17. Ahmad R, Xie L, Pyle M, Suarez MF, Broger T, Steinberg D, et al. A rapid triage test for active pulmonary tuberculosis in adult patients with persistent cough. Sci Transl Med. 2019;11(515).

18. Luo J, Zhang M, Yan B, Li F, Guan S, Chang K, et al. Diagnostic performance of plasma cytokine biosignature combination and MCP-1 as individual biomarkers for differentiating stages Mycobacterium tuberculosis infection. The Journal of infection. 2019;78(4):281–91.

19. Suarez I, Rohr S, Stecher M, Lehmann C, Winter S, Jung N, et al. Plasma interferon-gamma-inducible protein 10 (IP-10) levels correlate with disease severity and paradoxical reactions in extrapulmonary tuberculosis. Infection. 2021;49(3):437–45.

20. Pedersen JL, Barry SE, Bokil NJ, Ellis M, Yang Y, Guan G, et al. High sensitivity and specificity of a 5-analyte protein and microRNA biosignature for identification of active tuberculosis. Clin Transl Immunology. 2021;10(6):e1298.

21. Sampath P, Rajamanickam A, Thiruvengadam K, Natarajan AP, Hissar S, Dhanapal M, et al. Cytokine upsurge among drug-resistant tuberculosis endorse the signatures of hyper inflammation and disease severity. Sci Rep. 2023;13(1):785.

22. Álvez MB, Bergström S, Kenrick J, Johansson E, Åberg M, Akyildiz M, et al. A human pan-disease blood atlas of the circulating proteome. Science. 2025;390(6779):eadx2678.

23. Sun BB, Chiou J, Traylor M, Benner C, Hsu Y-H, Richardson TG, et al. Plasma proteomic associations with genetics and health in the UK Biobank. Nature. 2023;622(7982):329–38.

24. Kumar NP, Moideen K, Banurekha VV, Nair D, Babu S. Plasma Proinflammatory Cytokines Are Markers of Disease Severity and Bacterial Burden in Pulmonary Tuberculosis. Open Forum Infect Dis. 2019;6(7):ofz257.

25. WHO. High-priority target product profiles for new tuberculosis diagnostics Report of a consensus meeting. 2014.

26. WHO. Target product profile for tuberculosis diagnosis and detection of drug resistance. 2024.

27. Sutherland JS, van der Spuy G, Gindeh A, Thuong NTT, Namuganga A, Owolabi O, et al. Diagnostic Accuracy of the Cepheid 3-gene Host Response Fingerstick Blood Test in a Prospective, Multi-site Study: Interim Results. Clin Infect Dis. 2022;74(12):2136–41.

28. Gupta-Wright A, Ha H, Abdulgadar S, Crowder R, Emmanuel J, Mukwatamundu J, et al. Evaluation of the Xpert MTB Host Response assay for the triage of patients with presumed pulmonary tuberculosis: a prospective diagnostic accuracy study in Viet Nam, India, the Philippines, Uganda, and South Africa. Lancet Glob Health. 2024;12(2):e226–e34.

29. Sutherland JS, Mendy J, Gindeh A, Walzl G, Togun T, Owolabi O, et al. Use of lateral flow assays to determine IP-10 and CCL4 levels in pleural effusions and whole blood for TB diagnosis. Tuberculosis (Edinb). 2016;96:31–6.

30. Folkesson E, Fröberg G, Sundling C, Schön T, Södersten E, Bruchfeld J. Improved detection of extrapulmonary and paucibacillary pulmonary tuberculosis by Xpert MTB host response in a tuberculosis low-endemic, high-resource setting. The Journal of Infectious Diseases. 2025;232(1):e78–e88.

31. Chegou NN, Sutherland JS, Malherbe S, Crampin AC, Corstjens PL, Geluk A, et al. Diagnostic performance of a seven-marker serum protein biosignature for the diagnosis of active TB disease in African primary healthcare clinic attendees with signs and symptoms suggestive of TB. Thorax. 2016;71(9):785–94.

32. Mutavhatsindi H, van der Spuy GD, Malherbe ST, Sutherland JS, Geluk A, Mayanja-Kizza H, et al. Validation and Optimization of Host Immunological Bio-Signatures for a Point-of-Care Test for TB Disease. Front Immunol. 2021;12:607827.

33. De Groote MA, Sterling DG, Hraha T, Russell TM, Green LS, Wall K, et al. Discovery and Validation of a Six-Marker Serum Protein Signature for the Diagnosis of Active Pulmonary Tuberculosis. Journal of clinical microbiology. 2017;55(10):3057–71.

34. Zhao Y, Yang X, Zhang X, Yu Q, Zhao P, Wang J, et al. IP-10 and RANTES as biomarkers for pulmonary tuberculosis diagnosis and monitoring. Tuberculosis (Edinb). 2018;111:45–53.

35. Henno LT, Storjord E, Christiansen D, Bergseth G, Ludviksen JK, Fure H, et al. Effect of the anticoagulant, storage time and temperature of blood samples on the concentrations of 27 multiplex assayed cytokines - Consequences for defining reference values in healthy humans. Cytokine. 2017;97:86–95.

36. Patil R, Shukre S, Paranjape R, Thakar M. Heparin and EDTA anticoagulants differentially affect the plasma cytokine levels in humans. Scand J Clin Lab Invest. 2013;73(5):452–5.

37. Krishnan VV, Ravindran R, Wun T, Luciw PA, Khan IH, Janatpour K. Multiplexed measurements of immunomodulator levels in peripheral blood of healthy subjects: Effects of analytical variables based on anticoagulants, age, and gender. Cytometry B Clin Cytom. 2014;86(6):426–35.

38. Biancotto A, Feng X, Langweiler M, Young NS, McCoy JP. Effect of anticoagulants on multiplexed measurement of cytokine/chemokines in healthy subjects. Cytokine. 2012;60(2):438–46.

39. Assarsson E, Lundberg M, Holmquist G, Bjorkesten J, Thorsen SB, Ekman D, et al. Homogenous 96-plex PEA immunoassay exhibiting high sensitivity, specificity, and excellent scalability. PloS one. 2014;9(4):e95192.

40. Kursa MB, Rudnicki, W. R.. Feature Selection with the Boruta Package. Journal of Statistical Software, 36(11), 1–13. 2010.

41. Fonti V, Belitser, E. Feature selection using lasso. VU Amsterdam research paper in business analytics 30 (2017): 1–25. 2017.

42. Yi M, Nissley DV, McCormick F, Stephens RM. ssGSEA score-based Ras dependency indexes derived from gene expression data reveal potential Ras addiction mechanisms with possible clinical implications. Sci Rep. 2020;10(1):10258.

