## Supplemental material for "Multicountry validation of a proteomic host-response signature associated with tuberculosis disease severity"

***Supplementary Table 1.*** Demographic and clinical information of study participants from Sweden

| Characteristics | | TB disease (n=30) | TB infection (n=63) | non-TB disease (n= 26) | IGRA– controls (n=17) | Total (n=136) | *P value* |
|---|---|---|---|---|---|---|---|
| **Age, y, median (range)** | | 38 (18-77) | 34 (20-71) | 59 (23-79) | 38 (23-70) | 37 (18-79) | *p=0.004*[1] |
| **Females, n (%)** | | 16 (53) | 32 (51) | 11 (42) | 5 (29) | 64 (47) | *p=0.361*[2] |
| **Region of Origin** | W Europe | 10 (33) | 6 (10) | 6 (23) | 5 (28) | 27 (20) | *p=0.153*[2] |
| | E Europe | 2 (7) | 7 (11) | 2 (8) | 0 | 11 (8) | |
| | Africa | 11 (37) | 22 (35) | 7 (27) | 5 (29) | 45 (33) | |
| | Asia | 5 (17) | 25 (40) | 11 (42) | 6 (35) | 47 (35) | |
| | South America | 2 (7) | 3 (5) | 0 | 1 (6) | 6 (4) | |
| **IGRA-results** | Positive | 20 (67) | 62 (98) | 19 (73) | 0 | 101 (74) | *p<0.001*[2] |
| | Negative | 2 (7) | 0 | 6 (23) | 17 (94) | 25 (18) | |
| | N/A | 8 (27) | 1 (2) | 1 (4) | 0 (0) | 10 (7) | |
| **Specimen for TB diagnosis[b] (all TBD=30) n (%)** | Sputum | 12 (42) | - | - | - | | NA |
| | Induced Sputum | 0 | - | - | - | | |
| | BAL/BS/pleural fluid | 5 (17) | - | - | - | | |
| | Other sample | 13 (43) | - | - | - | | |
| **Smear results (PTB =17)** | Positive | 7 (41) | 0 | 0 | 0 | | NA |
| | Negative | 10 (59) | 9 (13) | 22 (85) | 3 (17) | | |
| | Not available | 0 | 53 (87) | 4 (15) | 15 (83) | | |
| **Smear grade (PTB =17) n (%)** | Negative | 10 (60) | - | - | - | | NA |
| | + | 0 | - | - | - | | |
| | ++ | 1 (7) | - | - | - | | |
| | +++ | 5 (33) | - | - | - | | |
| **DNA PCR[b] (all TBD=30) n (%)** | Positive | 18 (50) | 0 | 0 | 3 (17) | | NA |
| | Negative | 12 (40) | 10 (17) | 22 (85) | 0 | | |
| | Not available | 0 | 50 (83) | 4 (15) | 15 (83) | | |
| **Mtb culture[c], n (%)** | Positive | 28 (96) | 0 | 0 | 3 (17) | | NA |
| | Negative | 2 (4) | 10 (17) | 23 (89) | 0 | | |
| | Not available | 0 | 50 (83) | 3 (11) | 15 (83) | | |
| **Comorbidities[d], n (%)** | | 14 (47) | 14 (22) | 18 (69) | 5 (29) | 51 (38) | *p=0.110*[2] |
| **Immunosuppression[e], n (%)** | | 5 (17) | 5 (8) | 3 (12) | 0 (0) | 13 (10) | *p=0.300*[2] |
| **Invasive investigation[f], n (%)** | | 21 (72) | - | 5 (46) | - | 26 (19) | |
| **General symptoms[g], n (%)** | | 15 (50) | - | 7 (33) | - | 22 (16) | |

*non TB diagnosis: Respiratory tract infection (13), Post TB lung changes (4), Other lung disease (4), Benign lymphadenopathy, bacterial osteomyelitis, lung cancer, primary biliary sclerosis, no diagnose – symptoms resolved.

[b]PCR-positive by BD Max or GeneXpert MTB/RIF: Sputum (9). Lymphnode aspirate (5). BAL/BS (4). Intraabdominal biopsy (2). Ventricular lavage (1). Three individuals positive in two different sample types.

[c]Culture negative TBD: 1 PCR+ abdominal TB. 1 PCR+ pulmonary TB

[d]Comorbidities; *TB disease:* Diabetes Mellitus (4), hypertension (4), chronic kidney disease, congestive heart failure, chronic obstructive lung disease, asthma, hypothyreosis, thyreoiditis, paroxysmal atrial fibrillation, chronic hepatitis B, polymyalgia rheumatica, reumathoid arthritis. *TB Infection*: Psoriasis (5),

Diabetes Mellitus (4), hypertension (4), Morbus Crohn (2), rheumatoid arthritis, chronic hepatitis B. *Non-TB disease:* Diabetes Mellitus (6), reumathological disease (3), hypertension (2), chronic kidney disease (2), hepatitis B, chronic obstructive lung disease, hypothyreosis, multiple myeloma, Schistosomiasis, thyreoiditis. *Controls*: Asthma, hypertension (3), ulcerative colitis.

[e]Immunosuppression: due to medical conditions and treatments. *TB disease:* Prednisolon (3) TNFα-blocker. Chronic kidney disease. Lymphoma. All individuals with TBD were HIV-negative. *TB Infection*: Methotrexat (2), TNFα-blocker. Chronic kidney disease (2) *Non-TB disease*: Prednisolon, Rituximab, Chronic Kidney Disease.

[f]Invasive investigation: Bronchoscopy, Surgical biopsy, Fine needle aspiration.

[g] Fever, night sweats, loss of appetite, weightloss

[1]Kruskal-Wallis

[2]Fischer exact test

***Supplementary Table 2.*** *Clinical characteristics of Swedish study participants with TB disease*

| Characteristics | | PTB n = 17 | EPTB n = 13 | All TBD n = 30 | *P value* |
|---|---|---|---|---|---|
| **Female** | N (%) | 9 (53) | 7 (54) | 16 (53) | *p=1.000*[3] |
| **Age** | median (range) | 54 (25-77) | 32 (18-61) | 38 (18-77) | *p=0.017*[4] |
| **CRP** | mg/L, ref < 3 | 23 (1-82) | 18 (1-80) | 22 (1-82) | *p=0.983*[5] |
| **ESR**[a] | mm, ref < 20 | 44 (5-107) | 49 (1-115) | 47 (1-115) | *p=0.796*[5] |
| **Hemoglobin** | g/L, ref >120(F)/>130(M) | 126 (103-155) | 129 (93-158) | 127 (93-158) | *p=0.601*[5] |
| **Leukocyte count** | $10^9$/L,' ref 4.4-10.0 | 6.4 (4.0-9.9) | 6.8 (3.5-16.3) | 6.8 (4.0-16.3) | *p=0.706*[5] |
| **Albumine**[b] | g/L, ref >38 | 35 (26-45) | 34 (28-41) | 35 (26-45) | *p=0.871*[5] |
| **General symptoms**[c] | Yes (%) | 10 (59) | 5 (39) | 15 (50) | *p=0.462*[3] |
| **Invasive investigations**[d] | Yes (%) | 8 (47) | 13 (100) | 21 (70) | *p=0.003*[3] |
| **Microbiology** | Sputum microscopy+ | 7 (41) | - | | |
| | Sputum PCR+ | 9 (53) | - | | |
| | Non-sputum[1] PCR + | 6 (35) | 4 (36) | | |
| | Sputum culture + | 12 (71) | - | | |
| | Non sputum[2] culture+ | 8 (53) | 10 (91) | | |

***Pulmonary TB (PTB)***: 11 pulmonary TB (including 2 with pleural involvement), 4 primary pulmonary TB, 1 TB pleuritis, 1 disseminated TB (lungs and abdomen). ***Extrapulmonary TB (EPTB)***: 8 lymph node TB, 5 Abdominal TB.

[a] missing value in two EPTB. [b] missing value in four EPTB. [c] Fever, night sweats, loss of appetite, weightloss. [d]Bronchoscopy, Surgical biopsy, Fine needle aspiration.

[1]Non-sputum sample type: *PTB*: bronchial secretion (2), BAL (2), lymph node aspirate, lymph node biopsy, VL. Some individuals have several positive samples. *EPTB*: lymph node aspirate (2), peritoneal biopsy, ileocecal biopsy.

[2]Non-sputum sample type: *PTB*: Bronchial secretion (4) lymphnode biopsy/aspirate (3), pleural fluid, BAL, VL. Some individuals have several positive samples. Four individuals were only positive in non-sputum samples. *EPTB*: Lymphnode aspirate (9), peritoneal biopsy, colon biopsy, feces.

[3]Fischer's exact test

[4]Mann-Whitney test

*Abbreviations:* BAL = bronchioalveolar lavage. ESR = Erythrocyte sedimentation rate. VL = ventricular lavage

***Supplementary Table 3.*** *Demographic and clinical information of study participants from Italy*

| Characteristics | | HD | Pulmonary TB | TBI | Non-TBD[A] | Total | P value |
|---|---|---|---|---|---|---|---|
| **N (%)** | | 20 (14.8) | 31 (23.0) | 57(42.2) | 27 (20.0) | 135 (100) | |
| **Age median (IQR)** | | 42 (38-51) | 37 (28-42) | 46 (30-56) | 56 (42-70) | 44 (36-56) | .000 * |
| **Female N (%)** | | 14 (70) | 11 (35.0) | 33 (58.0) | 19 (70.0) | 77 (57.0) | 0.26** |
| **Origin N (%)** | **West Europe** | 19 (95) | 9 (29) | 38 (67.0) | 23 (85.0) | 89 (66.0) | .001** |
| | **East Europe** | 1 (5) | 15 (49.0) | 10 (18.0) | 2 (7.0) | 28 (21.0) | |
| | **Asia** | 0 (0) | 3 (10.0) | 2 (3.0) | 1 (4.0) | 6 (4.0) | |
| | **Africa** | 0 (0) | 2 (6.0) | 4 (7.0) | 1(4.0) | 7 (5.0) | |
| | **South America** | 0 (0) | 2 (6.0) | 3 (5.0) | 0 (0) | 5 (4.0) | |
| **BCG-vaccinated N (%)** | | 4 (20.0) | 22 (71.0) | 19 (33.0) | 4 (15.0) | 49 (36.0) | .000** |
| **TST N (%)** | **Negative** | 11 (55.0) | 1 (3.0) | 0 (0) | 1 (4.0) | 13 (10.0) | .000** |
| | **Positive** | 0 (0) | 12 (39.0) | 7 (12.0) | 0 (0) | 19 (14.0) | |
| | **N/A** | 9 (45.0) | 18 (58.0) | 50 (88.0) | 26(96.0) | 103 (76.0) | |
| **QFT-PLUS results (N%)** | **Negative** | 9 (45) | 0 (0) | 0 (0) | 20 (74.0) | 29 (21.0) | .000** |
| | **Positive** | 0 (0) | 8 (26.0) | 51 (89.0) | 0 (0) | 59 (44.0) | |
| | **Undetermined** | 0 (0) | 0 (0) | 6 (11.0) | 1 (4.0) | 1 (1.0) | |
| | **N/A** | 11 (55) | 23 (74.0) | 0 (0) | 6 (22.0) | 46 (34.0) | |
| **Smear results N (%)** | **Negative** | 0 (0) | 5 (16.0) | 4 (7.0) | 27 (100.0) | 36 (27.0) | N/A |
| | **Positive** | 0 (0) | 25 (81.0) | 0 (0) | 0 (0) | 25 (18.0) | |
| | **N/A** | 20 (100) | 1(3.0) | 53 (93.0) | 0 (0) | 74 (55.0) | |
| **Specimen N (%)** | **Sputum** | 0 (0) | 29 (93.0) | 3 (5.0) | 27 (100.0) | 59 (43.0) | N/A |
| | **Induced sputum** | 0 (0) | 2 (7.0) | 0 (0) | 0 (0) | 2 (2.0) | |
| | **BAL** | 0 (0) | 0 (0) | 1 (2.0) | 0 (0) | 1 (1.0) | |
| | **N/A** | 20 (100) | 0 (0) | 53 (93.0) | 0 (0) | 73 (54.0) | |
| **Smear grade N (%)** | **Negative** | 0 (0) | 5 (16.0) | 0 (0) | 0 (0) | 2 (1.5) | N/A |
| | **+** | 0 (0) | 3 (10.0) | 0 (0) | 0 (0) | 3 (2.0) | |
| | **++** | 0 (0) | 7 (23.0) | 0 (0) | 0 (0) | 7 (5.0) | |
| | **+++** | 0 (0) | 9 (29) | 0 (0) | 0 (0) | 9 (7.0) | |
| | **++++** | 0 (0) | 2 (6.0) | 0 (0) | 0 (0) | 2 (1.5) | |
| | **N/A** | 20 (100) | 5 (16.0)[#] | 57 (100) | 27 (100) | 112 (83.0) | |
| **Molecular results (N%)** | **Negative** | 0 (0) | 0 (0) | 1 (3) | 2 (8.3) | 3 (2.2) | N/A |
| | **Positive** | 0 (0) | 29 (93.5) | 0 (0) | 0 (0) | 29 (21.4) | |
| | **N/A** | 20 (100) | 2 (6.5) | 56 (97) | 25 (91.7) | 103 (76.4) | |
| **Culture results (N%)** | **Negative** | 0 (0) | 0 (0) | 4 (7.0) | 27 (100.0) | 31 (23.0) | N/A |
| | **Positive** | 0 (0) | 31 (100) | 0 (0) | 0 (0) | 31 (23.0) | |
| | **N/A** | 20 (100) | 0 (0) | 53 (93.0) | 0 (0) | 74 (54.0) | |

**Abbreviations:** N/A not available; BCG Bacillus Calmette-Guerin vaccine; TBI Tuberculosis Infection; TB Tuberculosis; Non-TBD non-TB disease; HD: healthy donors.

[A] The non-TB disease cohort included individuals with *Mycobacterium avium* complex (MAC, n=11), bacterial pneumonia (n=11), cavitating pneumonia (n=3), and lung cancer (n=1).

[#] In five subjects, the smear was scored as positive, but the smear grading was not available.

*Kruskal-Wallis Test; ** all p values are calculated by Chi Square of Pearson Test

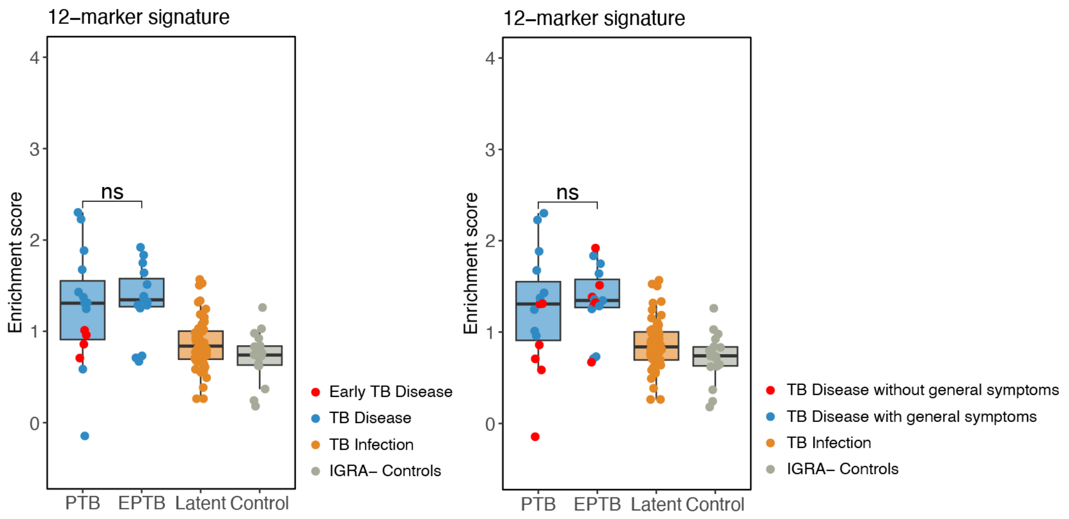


***Supplementary Figure 1.*** The enrichment score of the 12-marker signature in different subtypes of TB disease, including pulmonary TB and extrapulmonary TB. The left graph indicates individuals with Primary TB disease (found through contact tracing) indicated by red points. The right graph indicates individuals with general TB symptoms (blue points) or without general TB symptoms (red points). Statistics were evaluated using a Student's t-test with ns = p>0.05. **Abbreviations:** TB: tuberculosis; IGRA: Interferon-γ release assay; TBI: TB infection.

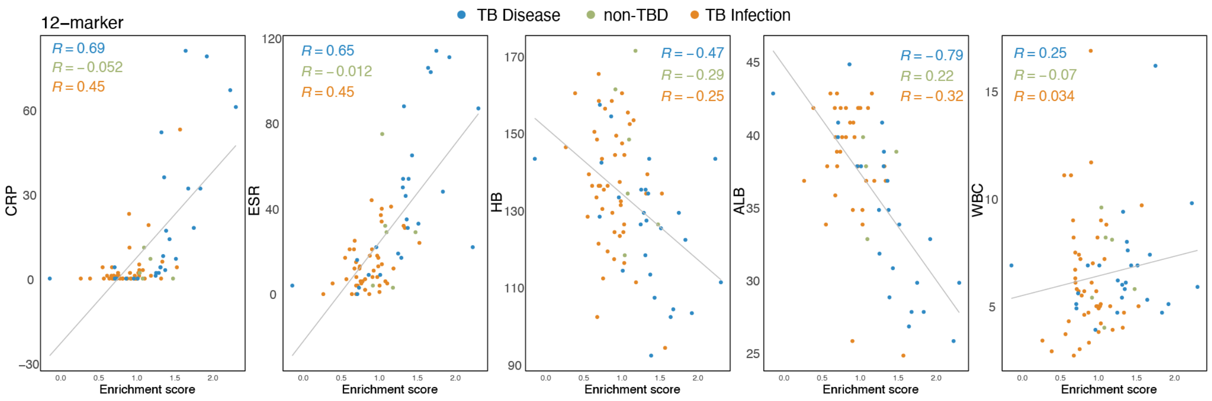


***Supplementary Figure 2.*** Correlation between enrichment scores from the 12-marker signature and various clinical blood chemistry markers, including C-reactive protein (CRP), Erythrocyte sedimentation rate (ESR), Hemoglobin (Hb), Albumin (Alb), and White blood cell count (WBC). The Pearson correlation coefficient was calculated on donors with TB disease (blue text), non-TB disease (green text), or TB infection (orange text) and is shown in the top of each graph.

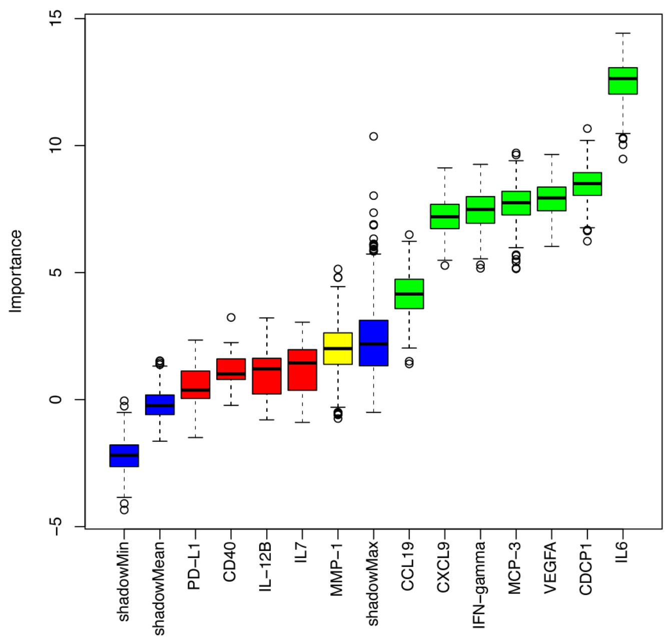


***Supplementary Figure 3.*** The importance level of each marker from the 12-protein signature in classifying TB disease versus TB infection, as calculated by Boruta**.** In Boruta, shadow features are generated by randomly permuting the values of each original feature in the dataset. The importance of both the original features and the shadow features is then calculated using a random forest algorithm. Boruta compares the importance of each original feature with the maximum importance of the corresponding shadow feature (i.e., shadowMax). Markers are colored based on their significance: confirmed important markers are shown in green, tentative markers in yellow, and rejected markers in red. Markers with higher importance than the shadowMax are considered relevant, while those with lower importance are rejected.

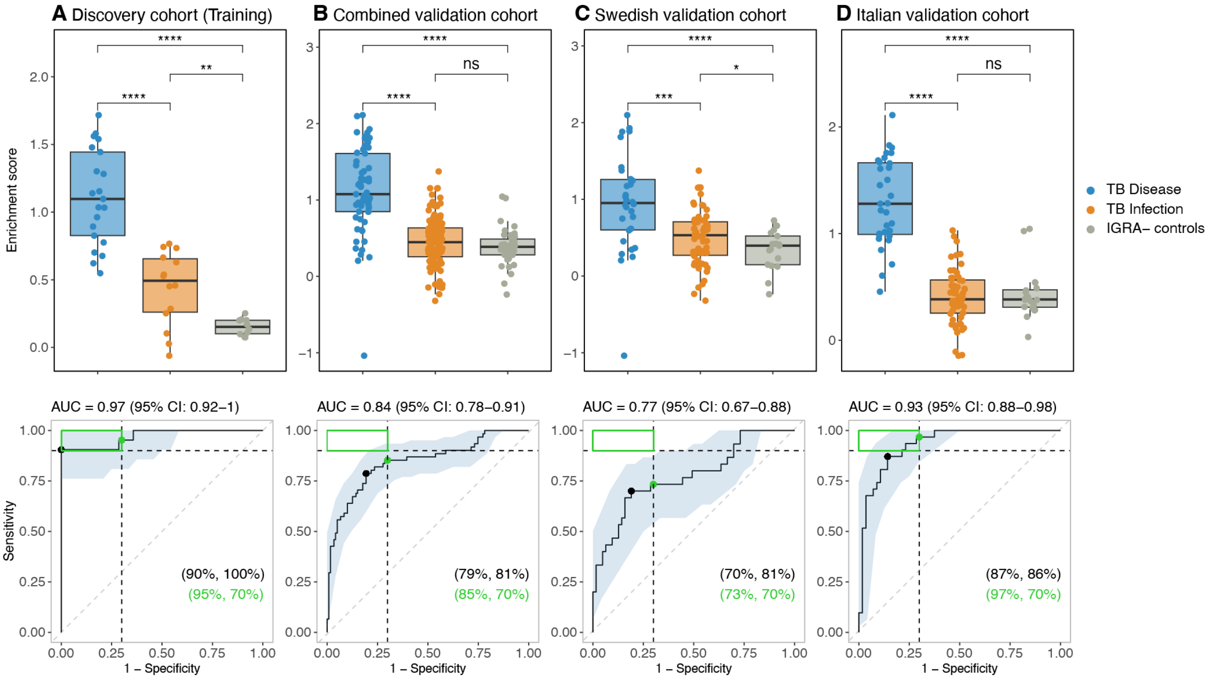


***Supplementary Figure 4.*** Accuracy of the 7-marker signature selected based on the Boruta output, for TB disease. The top row indicates single sample gene set enrichment analysis (ssGSEA) while the bottom row indicates ROC analysis, which was performed to classify TB disease versus TB infection. The shaded area indicates 95% confidence interval of the ROC curve, while the green box in the upper left corner indicates the minimum requirement for a triage test (sensitivity 90% and specificity 70%). The sensitivity at a fixed specificity of 70% for TB disease detection is indicated by a green point and the Youden index with a black point with percentages in the bottom right corner of the graph. The area under the ROC curve (AUC) with 95 % confidence interval is indicated above each graph. (**A**) Training cohort, (**B**) combined validation cohort, (**C**) Swedish validation cohort, and (**D**) Italian validation cohort. Statistics for the enrichment analysis were evaluated with Student's t-tests with ns=p>0.05, **p ≤0.01, ***p ≤0.001, ****p ≤0.0001. **Abbreviations:** TB: tuberculosis; IGRA: Interferon-γ release assay; ROC: Receiver operating characteristic; AUC: Area under curve.

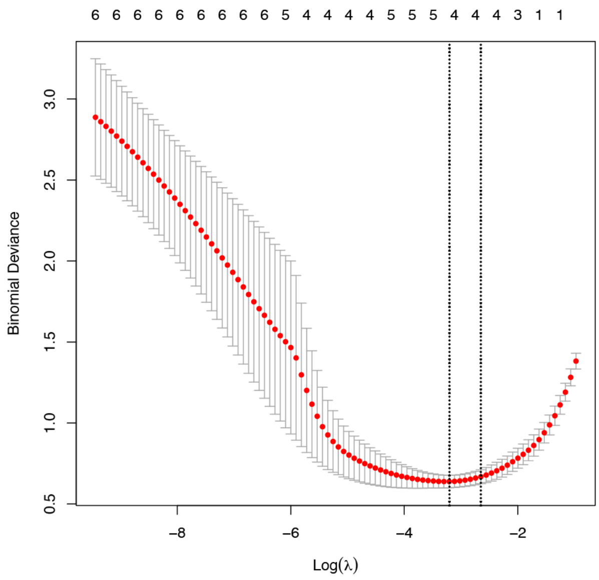


***Supplementary Figure 5.*** Binomial deviance for each combination of the 6-marker signature selected by Boruta, calculated using LASSO regression. A combination of 4 markers out of 6 markers had the lowest binomial deviance.

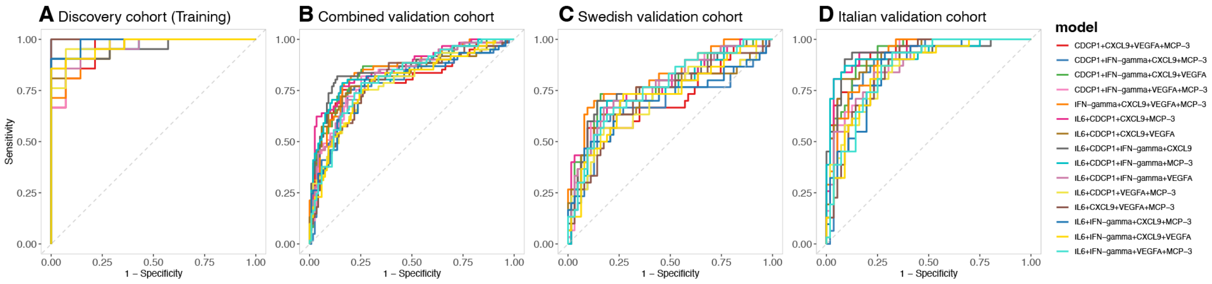


***Supplementary Figure 6.*** ROC curves for all combinations of 4 markers out of 6 markers, selected based on the Boruta output. Each color indicates a combination of 4 markers. (**A**) Training cohort, (**B**) combined validation cohort, (**C**) Swedish validation cohort, and (**D**) Italian validation cohort. **Abbreviations:** ROC: Receiver operating characteristic.

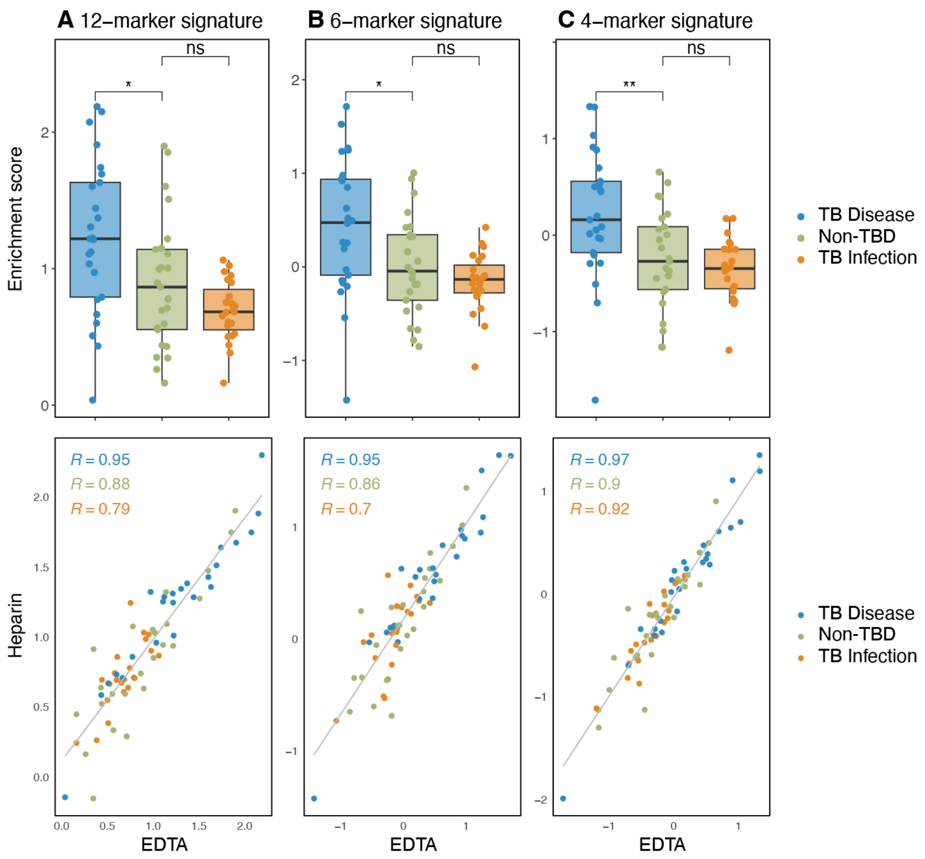


***Supplementary Figure 7.*** Anticoagulant effects on the performance of the 12-marker (**A**), 6-marker (**B**) and 4-marker (**C**) signatures. The top row indicates the enrichment scores in EDTA samples from the Swedish cohort (n=73). The bottom row indicates the Pearson correlation between enrichment scores for heparin and EDTA for the Swedish samples with TB disease (blue, n=25), non-TBD (green, n=24) and TB infection (orange, n=22) indicated and calculated separately. Statistics for the enrichment analysis were calculated using a Student's t-test with *p≤0.05, **p≤0.01.

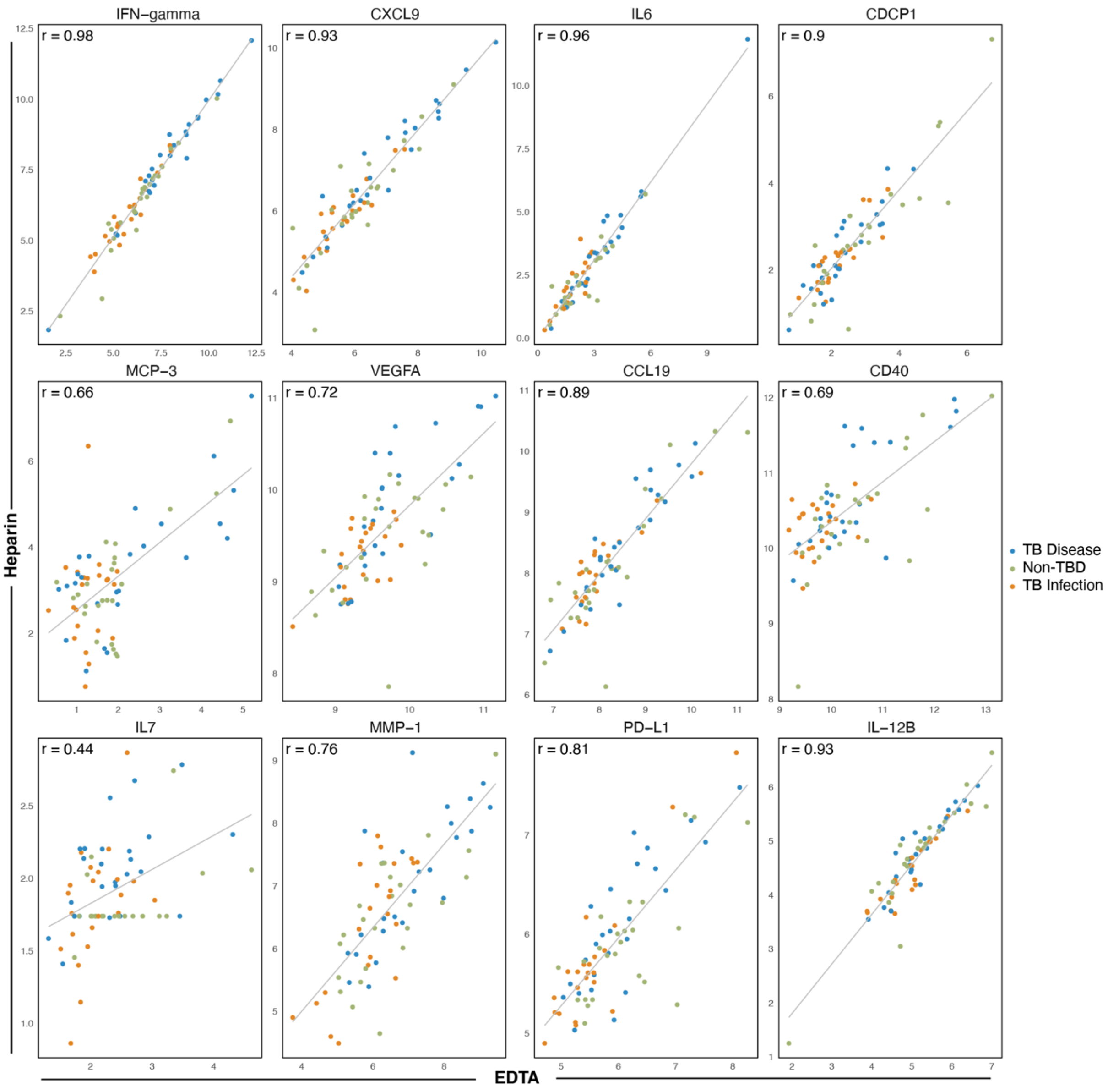


***Supplementary Figure 8.*** Pearson correlation between the NPX values from heparin (Y-axis) and EDTA (X-axis) samples for each individual protein included in the 12-marker signature (n=71 donors).

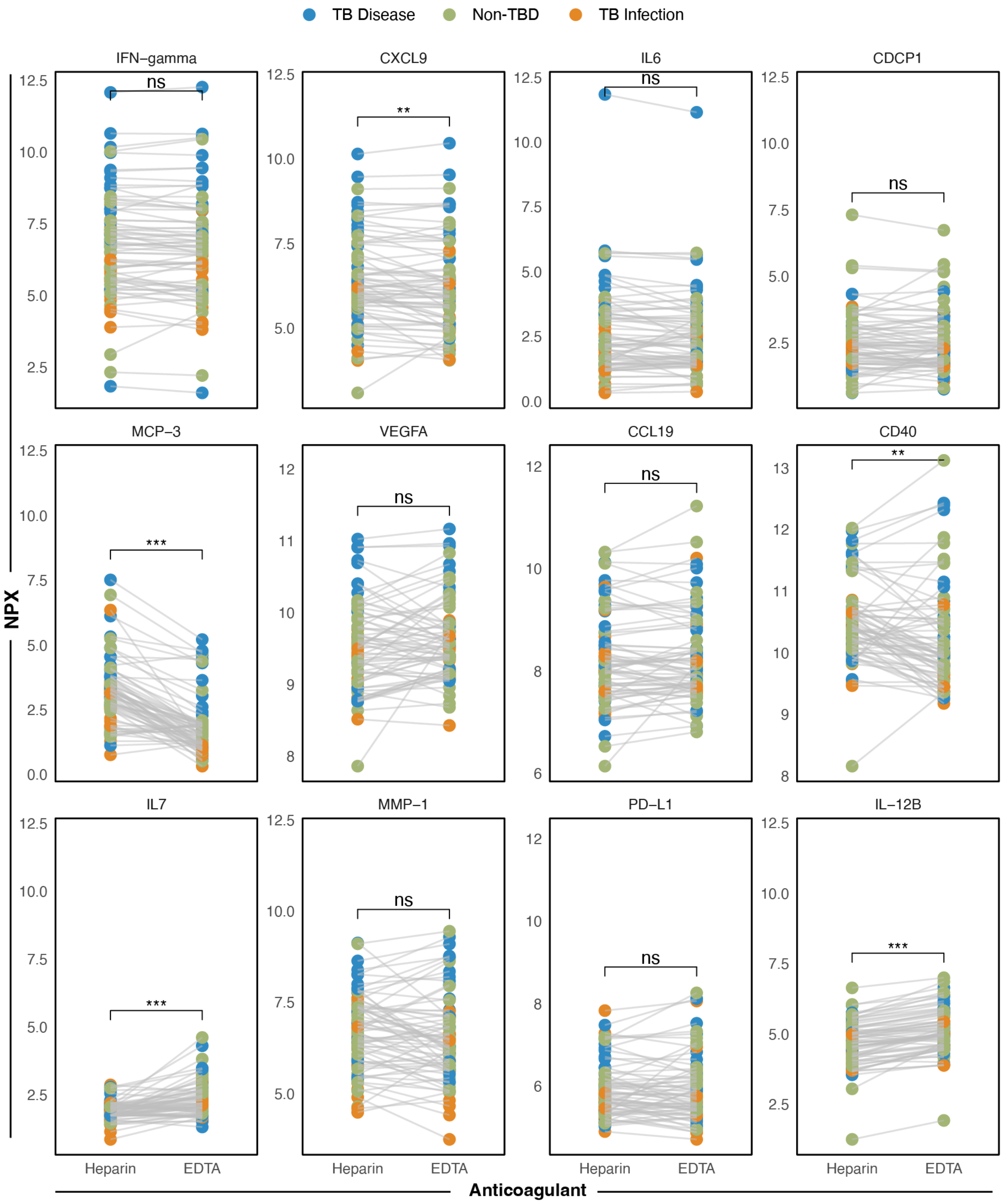


***Supplementary Figure 9.*** Comparison between NPX values for the individual proteins of the 12-marker signature. Donors with TB disease are indicated with blue points, non-TB disease with green points and TB infection with orange points (total n=71). Statistics were evaluated on pooled groups using paired t-tests with ns p>0.05, *p≤0.05, **p≤0.01, ***p≤0.001.

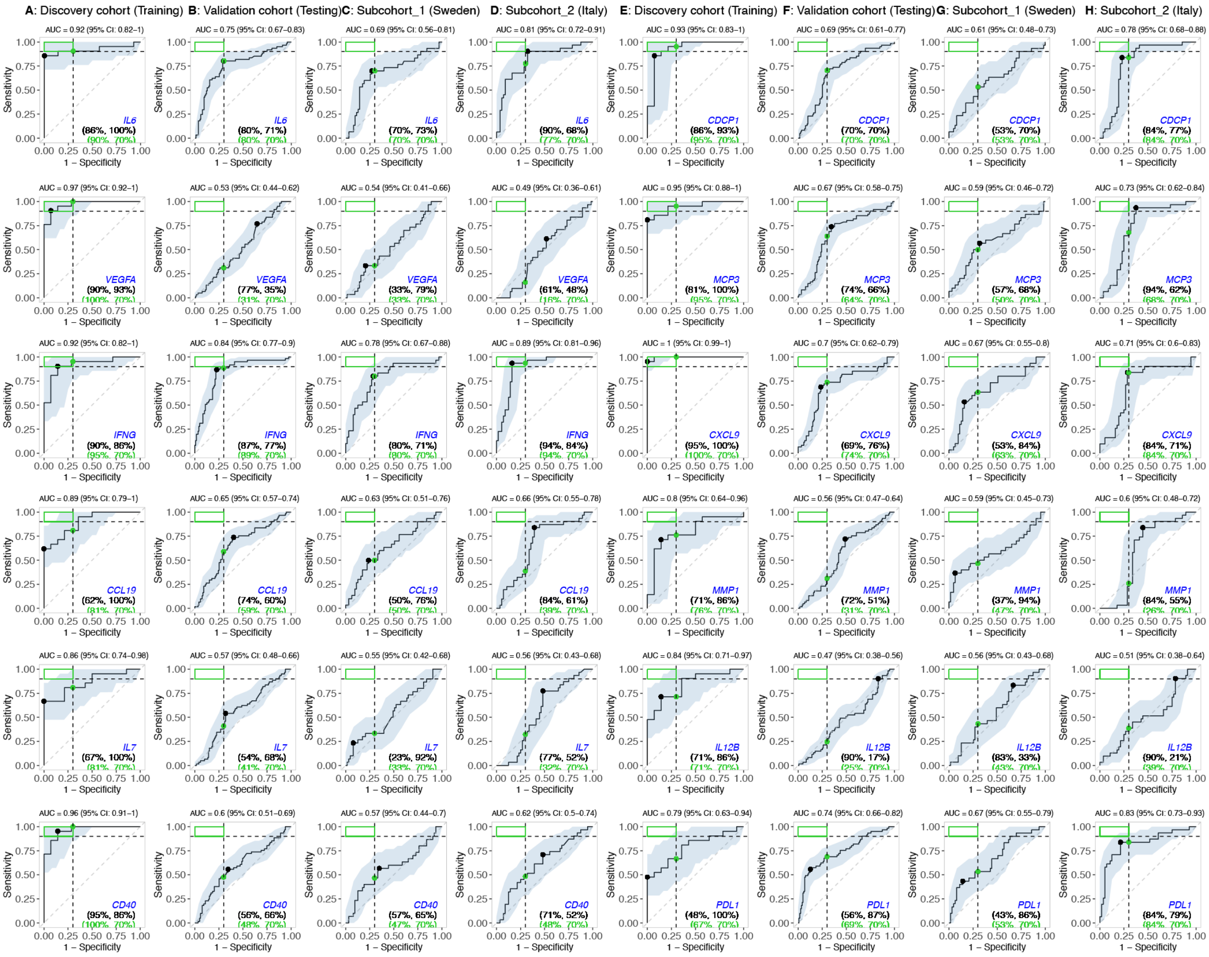


***Supplementary Figure 10.*** ROC analysis for each protein (indicated in blue in the plots) of the 12-marker signature for classifying TB disease versus TB infection. The shaded area indicates 95% confidence interval of the ROC curve while the green box in the upper left corner indicates the minimum requirement for a triage test (sensitivity: 90%, specificity: 70%). The sensitivity at a fixed specificity of 70% for TB disease detection is indicated by a green point and the Youden index with a black point with percentages in the bottom right corner of the graph. The area under the ROC curve (AUC) with 95 % confidence interval is indicated above each graph. (**A, E**) Training cohort, (**B, F**) combined validation cohort, (**C, G**) Swedish validation cohort, and (**D, H**) Italian validation cohort. **Abbreviations:** TB: tuberculosis; ROC: Receiver operating characteristic; AUC: Area under curve.

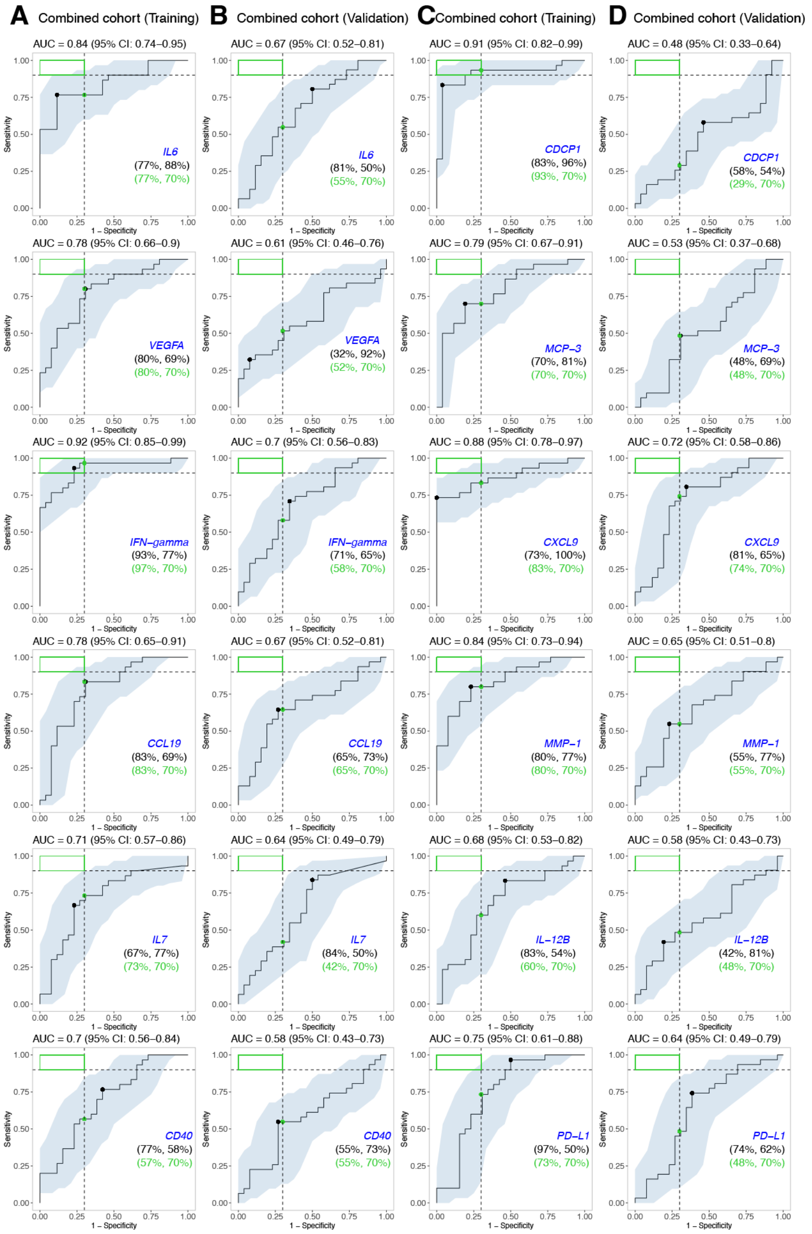


***Supplementary Figure 11.*** ROC analysis for each protein (indicated in blue in the plots) of the 12-marker signature for classifying TB disease versus non-TB disease. The shaded area indicates 95% confidence interval of the ROC curve while the green box in the upper left corner indicates the minimum requirement for a triage test (sensitivity: 90%, specificity: 70%). The sensitivity at a fixed specificity of 70% for TB disease detection is indicated by a green point and the Youden index with a black point with percentages in the bottom right corner of the graph. The area under the ROC curve (AUC) with 95 % confidence interval is indicated above each graph. (**A, C**) Training cohort, (**B, D**) validation cohort. **Abbreviations:** TB: tuberculosis; ROC: Receiver operating characteristic; AUC: Area under curve.

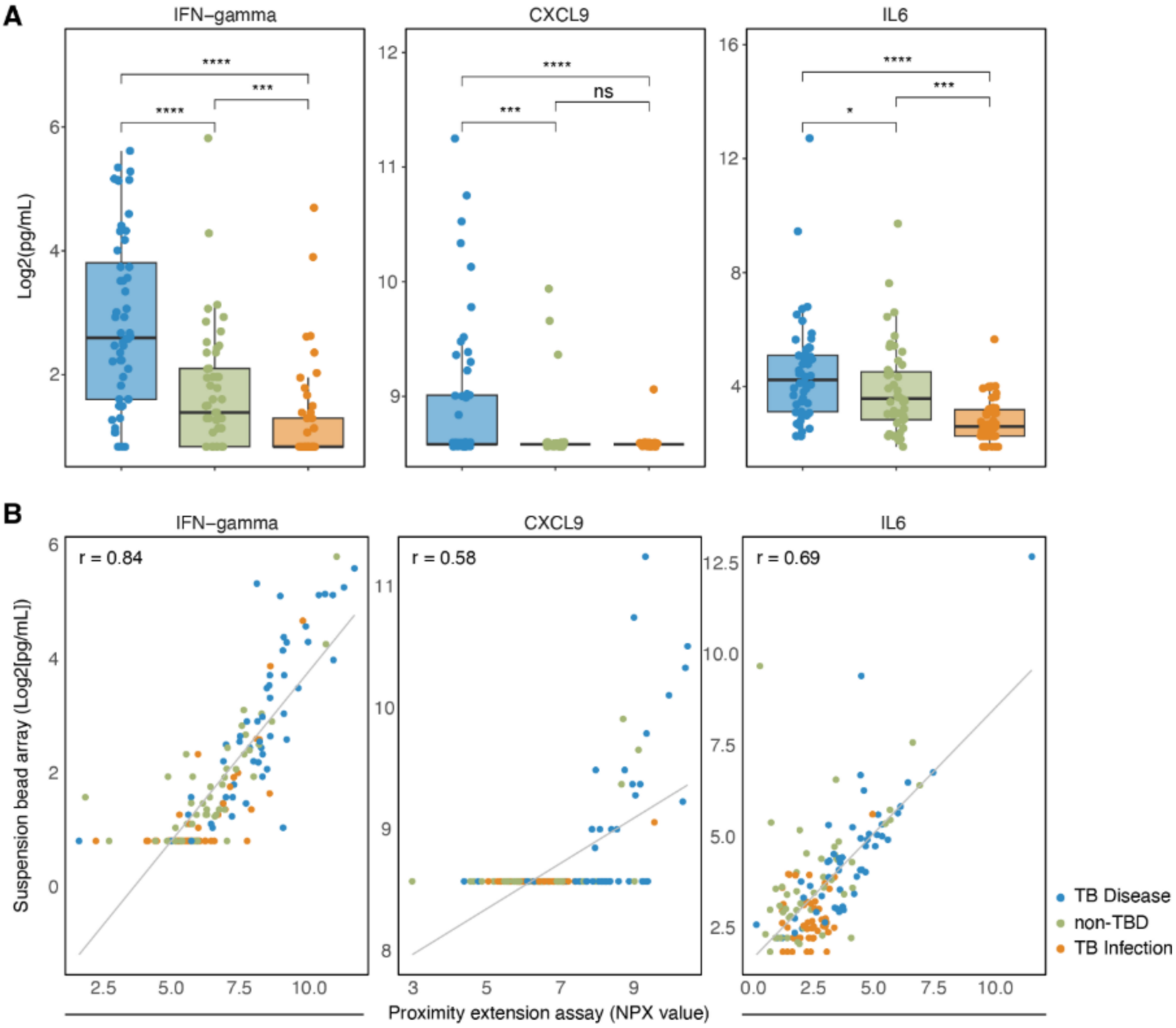


***Supplementary Figure 12:*** Cytokine levels quantified by suspension bead array and correlation with proximity-extension assay. (**A**) IFN-gamma, CXCL9, and IL6 levels were quantified by suspension bead array (Luminex or Legendplex) in heparin plasma from a subset of donors with TB disease (blue, n=52), non-TBD (green, n=50), and TB infection (orange, n=51). No assay for CDCP1 could be procured on the Luminex or Legendplex platforms. Cytokine levels were compared between groups using Student's t-tests with ns = p>0.05, *p<0.05, **p<0.01, ***p<0.001, ****p<0.0001. (**B**) Pearson correlation (r) was calculated between Log2 transformed suspension bead array-derived cytokine concentrations and proximity extension assay (Olink) normalized protein expression (NPX) values.
